# Population Differences in the Epidemiology, Phenotype, and Genetics of Hirschsprung Disease in the United States

**DOI:** 10.64898/2026.08.10.26360052

**Authors:** Mingzhou Fu, Hanna E Berk-Rauch, Monica Erazo, Sumantra Chatterjee, Aravinda Chakravarti

## Abstract

**Importance:** Understanding population differences in epidemiology, clinical presentation, and genetic architecture remains a major challenge for all rare genetic disorders. Hirschsprung disease (HSCR), despite being the commonest cause of neonatal intestinal obstruction, has been poorly studied with respect to its significant heterogeneity across U.S. populations.

**Objective:** To characterize self-identified race and ethnicity differences in HSCR incidence, clinical presentation, and genetic architecture in the United States from diverse data sources.

**Design, Setting, and Participants:** We used retrospective, population-based surveillance data from 3 independent US wide sources – (1) The National Birth Defects Prevention Network (NBDPN; 1996–2010), (2) aggregated electronic health record data from Epic COSMOS (1997–2025), and (3) individual-level clinical and genomic data from the Hirschsprung Disease Research Collaborative (HDRC; 2011–2025). Statistical analyses of incident HSCR cases identified at birth, across time and geography, in conjunction with clinical phenotypes and genome sequences from unrelated HDRC probands were performed to characterize epidemiologic, phenotypic and genetic heterogeneity in HSCR.

**Exposures:** HSCR cases were identified based on standardized clinical diagnostic criteria, primarily rectal biopsy with histopathologic confirmation of aganglionosis. The disease was defined using ICD-9-CM code 751.3, CDC/BPA code 751.30–751.34. and ICD-10-CM code Q43.1. Patients were classified by self-identified race and ethnicity (SIRE), with primary comparisons conducted between non-Hispanic Blacks/African Americans (Blacks) and non-Hispanic Whites (Whites).

**Main Outcomes and Measures:** HSCR incidence and the frequency of clinical features were estimated overall and by SIRE. We also estimated the individual and total genetic burden of rare pathogenic coding variants and common noncoding regulatory variants at established HSCR genes by population.

**Results:** Overall HSCR incidence in the U.S. was 2.04 per 10,000 live births (95% CI, 1.99–2.09) as previously estimated. We show, Blacks have the highest HSCR incidence (2.83–3.13 per 10 000 live births), in comparison to Whites (1.89–2.02) and Asians (1.54–1.98), a difference not previously ascertained from previous smaller cohorts from limited geographical regions. This difference persists across surveillance times and geography. This incidence difference from NBDPN is consistent with Epic COSMOS, a nation-wide, independent hospital-based data source. Clinically, Blacks are more likely to present with isolated HSCR and with milder manifestations at birth, including chronic severe constipation (CSC). Genetically, the burden of pathogenic coding variants did not differ between Blacks and Whites. However, Blacks had a significantly higher enrichment of two non-coding regulatory variants (rs199582499 and rs28735659) at the *SOX10* gene locus, as compared with Whites.

**Conclusions and Relevance:** This study demonstrates, for the first time, that Black HSCR patients in the U.S. have a higher incidence accompanied by milder clinical presentation and distinct noncoding regulatory *SOX10* variants as compared to White patients. Nevertheless, Blacks are severely under-represented in U.S. studies of HSCR leading to significant health disparities in their care and management.

**Key Points:** *Question:* Are there population differences in the incidence, clinical presentation, and genetic architecture of Hirschsprung disease in the United States?

*Findings:* In this population-based study integrating national surveillance data, electronic health records, and results of whole-genome sequencing, Black patients showed the highest incidence of HSCR but with milder clinical presentation than Whites. Genetically, the major difference in Blacks is the unique association with noncoding regulatory variation at the *SOX10* gene not observed in Whites.

*Meaning:* These findings uncover significant population differences in incidence and clinical presentation, but limited qualitative but greater quantitative genetic differences, in Hirschsprung disease. Beyond being useful for understanding HSCR natural history and for genetic counseling, it begs for research on the causes of the variable frequency and presentations across populations.

## Introduction

Hirschsprung disease (HSCR), also known as aganglionic megacolon, is a congenital disorder of the enteric nervous system (ENS) caused by failure of enteric neural crest cell–derived cells (ENCDCs) to fully colonize the gastrointestinal tract during embryonic development.^1,2^ The disease is associated with the absence of ENCDCs or ganglion cells in both the myenteric and submucosal plexuses resulting in the commonest functional intestinal obstruction in neonates, usually affecting the distal colon.^3–6^ Clinically, HSCR presents in infancy with delayed passage of meconium, abdominal distension, feeding intolerance, and severe constipation and typically requires surgical intervention.^4,7^ The leading diagnostic symptom in 60% of cases is the inability to pass meconium in the first 2 days of life, but can be absent or overlooked, and 30% of patients are diagnosed later in infancy, sometimes (<10%) in older children and adults with chronic severe constipation.^8^ Despite surgery, 30-50% of patients carry substantial, long-term consequences with some studies showing reductions in quality of life.^9–12^ Since HSCR is not an uncommon disorder in pediatric populations and displays highly significant phenotypic variation at birth,^8^ we consider it important to revisit its epidemiology and clinical presentation to estimate its clinical burden today.

One of the first epidemiological studies of HSCR frequency was by Goldberg who estimated its overall incidence at approximately 1.86 per 10,000 live births.^13^ However, subsequent studies across the world have reported rates that vary considerably across populations and geographic regions – ranging from 1.1-2.3 in Europe, ^14–20^ from 1.8-2.2 in East Asia,^21–24^ and 3.3 in the Arabian Peninsula, all per 10,000 births.^25^ In the U.S., earlier population-based studies suggested modest population differences in HSCR incidence, including higher incidence among non-White populations, particularly in Asians.^13,26^ However, many of these estimates were derived from older birth cohorts, limited geographic regions, or relatively small registries. More recent data from a large California birth-defects registry have reported substantially higher HSCR incidence among Black/African American patients as compared with other groups,^27^ raising questions about whether prior estimates underestimated population-specific risk. Although HSCR has very high heritability (>85%)^5^ for an assumed multifactorial disorder, one cannot disprove that environmental effects are contributory. Earlier studies have suggested specific risk factors, such as maternal hyperthermia,^28^ but these are unproven.^29^ More recently, the contributions of vitamin A deficiency, ibuprofen, mycophenolate and diet and exercise have been suggested but these are also unproven.^30–33^ Since environmental risk factors and their exposures can change dramatically across time, we consider it important to systematically reevaluate the epidemiology of HSCR using multiple nationwide sources of the U.S. across populations using contemporary, multi-state surveillance data over an extended period.

HSCR is clinically a highly heterogeneous male-biased (sex ratio 3.6) developmental anomaly characterized by aganglionic segment length (short, long and total colonic aganglionosis), a high frequency of associated congenital anomalies including Down syndrome, and familiality. ^8^ While most cases are diagnosed in the neonatal period with rectal biopsy and Acetylcholinesterase (AChE) staining, milder presentations are associated with delayed diagnosis of HSCR, ^34,35^ or a diagnosis of chronic severe constipation (CSC).^36,37^ The phenotypic diversity of HSCR presentation is also associated with disease severity and these features may impact their ascertainment in surveys and, thus, their reported incidence. In addition, it is unknown whether such clinical presentations vary across U.S. population.

Genetically, HSCR is a multifactorial or complex genetic disorder with contributions from both rare coding variants and common noncoding regulatory variants. To date, more than 24 genes have been shown to harbor pathogenic variants associated with HSCR in total explaining 62% of its population attributable risk (PAR).^38^ Importantly, at least 10 of these genes are functionally united through a gene regulatory network (GRN) that modulates the expression of two major genes, *RET* and *EDNRB*, during ENS development.^39^ Unsurprisingly, ∼20-50% of HSCR’s PAR arises from these two major genes.^5,38^ Moreover, a set of 5 transcriptions factors (TF) control gene expression of *RET* and *EDNRB* and are HSCR genes. Of these, *SOX10* has a major role since pathogenic mutations in this TF leads to a dominant form of syndromic HSCR.^40^ Although much has been learnt regarding HSCR genetics, these studies have largely been conducted in European- and Asian-ancestry subjects at the near virtual exclusion of African-ancestry cases, ^38,41–49^ leaving a major gap in the field. It has generally been assumed that HSCR incidence is higher in Asians than Whites in the U.S.,^26^ but supporting evidence is limited and largely based on regional studies,^13,26,27^ underscoring the need for more comprehensive data and more precise incidence estimates.

To address this significant gap, we combined national population-based surveillance data with a deeply phenotyped, multicenter clinical-genetic cohort to examine population differences in HSCR incidence, clinical presentation, and its genetic architecture in the U.S. For comparisons and classifications, we use self-identified race and ethnicity (SIRE) categories as defined in OMB’s 1997 Directive No. 15.^50^ Using these datasets, we re-estimated HSCR incidence, investigated clinical phenotypic differences and explored the genetic risk profiles across SIRE, with particular focus on comparisons between non-Hispanic Black/African American (Black) and non-Hispanic White (White) patients.

## Materials and Methods

### Study Design and Data Sources

We conducted a retrospective, observational cohort study integrating population-based surveillance data, electronic health records data, and a multicenter, randomly ascertained HSCR research cohort to evaluate U.S. population differences of HSCR in its epidemiology, clinical presentation, and genetic architecture.

Epidemiologic analyses were conducted using two national, aggregate-level data sources, namely, the National Birth Defects Prevention Network (NBDPN) and Epic COSMOS.^51,52^

The NBDPN is a collaborative network of U.S. state-based birth-defects surveillance programs that collects population-level data using standardized case definitions, diagnostic confirmation practices, and SIRE- and state-specific birth counts.^52^ State-level birth-defect surveillance reports are released in 5-year reporting intervals, and both HSCR case counts and numbers of live-births, stratified by SIRE and birth state, are available on their website (https://nbdpn.org/birth-defects-data-tables-and-directory/). For this study, HSCR incidence data from 1996 through 2010 were used, corresponding to the period during which HSCR was consistently captured across participating states with the same SIRE categories based on standards from the Office of Management and Budget (OMB).^50^ HSCR cases were identified using ICD-9-CM code 751.3 or CDC/BPA codes 751.30–751.34, and population groups were reported as non-Hispanic White (White), non-Hispanic Black/African American (Black), non-Hispanic Asian or Pacific Islander (Asian), non-Hispanic American Indian or Alaska Native (Native), Hispanic, and Other/Unknown. Data from 1989–1995 were excluded because of inconsistent SIRE classification (Table S1). HSCR reporting was discontinued in NBDPN in 2014. NBDPN data were used as the primary source for HSCR incidence estimation because of its population-based surveillance design and relatively standardized reporting across states.

Epic COSMOS is a large electronic health record (EHR) aggregation platform containing de-identified data from more than 176 million patients across 191 U.S. health care facilities (https://cosmos.epic.com/).^51^ Aggregated-level data were accessed using SlicerDicer, a web-based query tool, operating in de-duplicated patient mode.^53^ Live births were identified by specifying “Diagnosis (All)” as “Livebirth”, and HSCR cases were identified by specifying “Diagnosis (All)” as “Hirschsprung disease” (ICD-10-CM: Q43.1). SIRE categories included non-Hispanic White (White), non-Hispanic Black/African American (Black), non-Hispanic Asian (Asian), non-Hispanic Native (Native: Native Hawaiian/Other Pacific Islander or American Indian/Alaska Native), and Hispanic/Latino. Epic COSMOS data from 1997 through 2025 were analyzed. Because Epic COSMOS reflects hospital-based clinical encounters with potential variation in diagnostic coding and ascertainment across institutions, it was used as a secondary data source to corroborate epidemiologic patterns, rather than as the primary source for analysis.

To contextualize U.S. HSCR incidence estimates within the global literature, a PubMed literature search was conducted using the terms “(Hirschsprung OR megacolon) AND (incidence OR prevalence)”, restricted to English-language publications from 1950 onwards – when the pathology-based HSCR diagnosis was established.^54^ A total of 361 unique articles were identified but, after title and abstract screening, 282 articles were excluded for not reporting HSCR incidence or birth prevalence, leaving 79 articles for full-text review. Among these, 16 studies were included as they: (1) explicitly reported HSCR incidence or at-birth prevalence; (2) were based on population-level surveillance or multi-center medical records; (3) had defined cohorts and study periods; and (4) provided explicit case or live-birth counts. These 16 studies represent populations from North America, Oceania, Europe, and East Asia. Of these, 9 studies with mid-study periods around 1996 were selected for analysis to match the NBDPN study periods, ensuring temporal consistency for comparison (Table S2). The Supplementary Methods detail the literature analysis.

Clinical phenotype and genetic analyses were performed using individual-level phenotype data from the Hirschsprung Disease Research Collaborative (HDRC), a national, multicenter research collaborative established in 2011 and led by Dr. Aravinda Chakravarti, with the coordinating center at NYU Grossman School of Medicine (IRB #i18-00033). The HDRC was designed to identify patients with HSCR at or near their initial presentation for pediatric surgical care and to systematically collect standardized demographic, clinical, phenotypic, and biological sample data to improve understanding of HSCR etiology, treatment, and outcomes (https://clinicaltrials.med.nyu.edu/clinicaltrial/605/hirschsprung-disease-research-collaborative/). Participants were recruited from 25 pediatric surgical centers and affiliated clinics across the United States, after written informed consent, and both affected individuals and their (usually) first-degree relatives were enrolled. Detailed clinical and phenotypic data were abstracted from medical records and stored in a secure REDCap database,^55^ while biospecimen and genomic data were managed through a LabVantage laboratory information management system (LIMS),^56^ under oversight by NYU Langone Health; access to individual-level identified or de-identified data requires institutional review board approval. As of July 2024 (2011-2024), the HDRC had enrolled 4,607 individuals with HSCR and their relatives, including more than 900 participants with biospecimens with DNA extracted, genotyped, or sequenced, making it the largest and most comprehensive collection of HSCR clinical and genetic data in North America (Table S3).

### Incidence Estimation and Comparison

Incidence was calculated as the number of live-born children diagnosed with HSCR before 2 years of age per 10,000 live births. Live-birth denominators were obtained from the corresponding surveillance systems. Ninety-five percent confidence intervals were calculated using the Murphy and Wolf method.^57^ Incidence was estimated both overall and stratified by SIRE, reporting time intervals and state.

For stratified analyses, all SIRE groups were standardized to OMB categories^50^ and restricted to non-Hispanic individuals. Hispanic ethnicity was analyzed as a separate category. Temporal analyses were conducted across pre-defined NBDPN’s reporting time intervals (1996–2000, 2001–2005, and 2006–2010). Geographic variation was assessed at the state level. Incidence rate ratios were calculated to compare HSCR incidence across populations,^58,59^ with incidence among White patients as the reference since they represent the largest numbers of births. Additionally, to assess the persistence of group differences in HSCR incidence in NBDPN, a multivariable regression analysis was performed by setting HSCR incidence as the response variable and setting SIRE group, time interval, longitude and latitude as explanatory variables, using Whites as the reference group. The R GLM package was used for the analysis.

### Clinical Phenotype Analysis (HDRC)

For this study, analyses were restricted to the 380 unrelated probands with a clinically confirmed diagnosis of HSCR as documented in their medical record rather than self-report. Family members, be they affected or unaffected, were excluded to avoid nonindependence of observations. Eligible probands met the following criteria: provision of informed consent; ascertainment during the study period; and a clinical diagnosis of HSCR documented in the medical record rather than by self-report. Probands who withdrew from the study were excluded.

### Population classification

SIRE information in the HDRC cohort was obtained primarily through self-report, using standardized questionnaires aligned with OMB categories,^50^ however, 21% had missing or unknown SIRE information. To compensate for this large degree of missing data, we used instead their estimated genetic ancestry, relative to the 1000 Genomes reference populations, using their whole genome sequence (WGS) data.^60^ Ancestry-inferred categories were assigned based on predefined ancestry proportion thresholds supported by prior population genetics studies. Thus, individuals were classified as non-Hispanic White (White) if European (EUR) ancestry was greater than 0.95 and East Asian (EAS) ancestry less than 0.05,^61,62^ non-Hispanic Asian (Asian) if eastern-Asian (EAS) ancestry was greater than 0.90,^63^ and non-Hispanic Black (Black) if African (AFR) ancestry was greater than or equal to 0.70 with EUR less than 0.30 and EAS less than 0.05.^61,64–66^ Individuals whose ancestry proportions did not meet these criteria were classified as other or undefined. Sensitivity analysis using 205 non-Hispanic patients with both SIRE and genetic ancestry showed high concordance – all inferred race labels were consistent with SIRE. Indeed, inferred genetic ancestry is more conservative and may remain undefined even when SIRE is reported, therefore minimizing misclassification (Figure S1). Final group classification followed a hierarchical approach: SIRE was used when available and genetic ancestry-inferred labels were used only when SIRE was missing or reported as unknown.

This led to a final cohort of 309 patients with defined population affiliations, reducing the unknown/missingness to 8%: of these, 237 were non-Hispanic Whites (Whites) and 37 were non-Hispanic Blacks (Blacks) and were used for downstream analyses (Figure S2). Other groups had too few samples for reliable statistical inference. The relative numbers of Whites and Blacks in HDRC was 6.4, comparable to the corresponding ratio in the U.S. general population, based on the 2020 U.S. Census (5.2)^67^ (P= 0.26). Thus, the population composition of the HDRC is broadly representative of the U.S. population with respect to the most frequent groups of Whites and Blacks.

### Phenotype and Clinical Comparisons

Phenotypes and clinical variables evaluated in this study included demographic characteristics (mortality status, age at diagnosis and sex), diagnostic methods (e.g., rectal biopsy and contrast enema), initial clinical manifestations at diagnosis, HSCR specific phenotypes (segment length, familiality, syndromic status), and all medical complications in the medical charts. Initial HSCR manifestations were abstracted from clinical notes and categorized into predefined symptom groups, including chronic severe constipation (CSC), abdominal distension, delayed passage of meconium, enterocolitis, and other presenting features. CSC was defined based on clinical documentation consistent with pediatric gastroenterology guidelines.^36,37^ Additionally, to aggregate HSCR-specific phenotypes, a HSCR severity score was calculated for individual patients as previously described.^68^ Briefly, individuals who were female, had longer segment length, had HSCR associated syndromes, and were from multiplex families had higher severity scores on each such factor, which were then summed for an overall score.

Phenotype comparisons of population groups were conducted using Fisher exact tests or χ² tests for categorical variables and Wilcoxon rank-sum tests for continuous variables, as appropriate. Odds ratios (OR), 95% confidence intervals of the OR (95%CI) and significance levels (P) were obtained using standard methods. All statistical tests were two-sided and an overall significance threshold of P < 0.05 was used.

### Genetic Analyses

#### Sample Preparation, Variant Calling and Prioritization

Genomic DNA was extracted from 2,366 blood samples and 117 saliva samples from HSCR probands and their relatives in the HDRC cohort using standard protocols, followed by quality control. DNA samples were properly aliquoted, plated and shipped to the Broad Institute for whole-genome sequencing (WGS), funded by the NIH Gabriella Miller Kids First Pediatric Research Program (project HD110884-01). Sequencing was performed on Illumina platforms using paired-end 150 base pair reads, targeting a mean genome-wide coverage of 30x.^69^ Reads were aligned to the human reference genome (hg38), and variants were called using HaplotypeCaller. Sample- and variant-level quality control were performed following GATK’s Best Practices.^70^

The interval-scattered, joint-called variant files (VCF.gz) were downloaded from the NIH’s secured data portal, and unrelated HDRC probands were extracted for downstream analyses. Additionally, following our laboratory’s standard data processing pipeline,^38,71^ ancestry estimation, sex inference and variant annotations were performed. Particularly, pathogenic coding variants (PVs) were defined and prioritized to include missense, loss of function (LoF) and insertion or deletion (INDEL) variants with predicted deleterious functional consequences. PVs were further restricted to rare variants (with a global allele frequency less than 1% in reference populations.^72^), and these rare PVs were used in downstream burden analyses.

Details of these procedures are described in the Supplementary Methods.

#### Rare Coding Pathogenic Variant Burden Analysis

Burden analyses of rare coding pathogenic variants (PVs) were performed for individual genes to compare HSCR-associated genetic risk between White and Black probands. Analyses focused on previously established 24 HSCR risk genes.^38^ For each gene, probands were classified as carriers or non-carriers based on the presence of at least one PV. Odds ratios and two-sided P values were calculated using Fisher’s exact tests to assess differences in PV carrier frequency between SIREs.

#### Definition of cis-regulatory elements (CREs) and their Variants

Candidate CREs were identified by defining chromatin accessibility data (DNase I hypersensitive sites (DHSs) of human embryonic colon tissues (doi:10.17989/ENCSR857AEB) from the ENCODE project,^74,75^ a development stage and tissue type associated with HSCR,^76,78^ focusing on the 24 HSCR risk genes. We defined the regulatory window for each CRE by restricting them to publicly available transcriptional topologically associating domains (TADs). ^73,77^ Common regulatory enhancer variants were defined and prioritized according to the following criteria. First, we chose common noncoding variants, defined as variants outside exonic coding regions in GENCODE data^79^ with a global allele frequency greater than 5% in at least one major ancestry group (non-Finnish European, African/African American, or East Asian) in the gnomAD v4 database. ^72^ Second, variants had to be located within candidate CREs. Separately, we also evaluated 10 experimentally validated HSCR functional enhancer variants at the *RET* gene locus.^80^ For each enhancer variant, population-specific allele frequencies observed in the HDRC probands were compared with the reference allele frequencies from African Americans and non-Finnish Europeans in the gnomAD v4 database using Fisher exact tests.

#### Haplotype comparisons at the *RET* locus

Because HSCR enhancer variants function synergistically,^39^ for the 10 experimentally verified enhancer variants at the *RET* locus, a haplotype comparison between groups was performed following previously published methods.^80^ Briefly, individual haplotypes of HDRC cases were estimated by phasing WGS data at the *RET* locus using Beagle^81^ and SHAPEIT5.^82^ Haplotype frequencies of the general population (control) were obtained separately for Blacks and Whites from the 1000 Genomes and GTEX databases (dbGaP study accession: phs000424.v2.p1).^60,83^ Specifically, control samples included 131 Black (61 ASW from 1000 Genomes and 70 self-reported Black/African Americans from GTEx) and 972 Whites (404 non-Finnish European individuals from 1000 Genomes and 568 self-reported White individuals from GTEx). Only haplotypes with a frequency greater than 1% in the group-specific control population were retained for analyses.

Haplotype containing the fewest risk enhancer variants (lowest-risk haplotype) was used as the reference. For each haplotype, the ratio of its frequency relative to the reference haplotype was computed separately for cases and controls. These ratios were then compared between cases and controls to estimate haplotype-specific risk ratios (odds ratios). Statistical significance was assessed using Fisher exact tests.

To understand the genetic relationship of the CRE variants by population, additionally, pairwise linkage disequilibrium (LD) among the variants was evaluated using phased 1000 Genomes reference data, using ASW samples for Blacks and CEU samples for Whites. Pairwise LD metrics, LD block structures, and population-specific haplotype frequencies were calculated and visualized using Haploview.^84^

#### Multi-test adjustments

Because multiple variants and genes were tested, we used the Bonferroni correction for all analysis.^85^

## Results

### Population Differences in HSCR Incidence

For consistency, HSCR incidence is reported per 10,000 live births across all analyses. Using data from NBDPN, the overall incidence of Hirschsprung disease (HSCR) in the U.S. is estimated at 2.04 (95% CI, 1.99–2.09) during the study period 1996-2010 (Figure 1A). When stratified by SIRE, HSCR incidence differed significantly across groups with Blacks exhibiting the highest incidence, with an estimate of 2.98 (95%CI: 2.83–3.13), as compared with 1.95 (95%CI: 1.89–2.02) among Whites and 1.75 (95%CI: 1.54–1.98) among Asians (Figure 1A). Thus, comparing the means of the three U.S. populations studied, Asians have the lowest incidence of HSCR, followed by Whites, with the highest rate being in Blacks. Comparisons of their 95% confidence limits shows some overlap of Whites and Asians (P= 0.48) but the incidence in Blacks is significantly 53% higher (P= 4.8 × 10^-41^) than Whites. This Black-White difference in HSCR incidence is consistent over time. Across successive surveillance time intervals, the incidence rate ratio comparing Black with White cases is remarkably constant, with estimates of 1.6, 1.5, and 1.5 during the periods 1996–2000, 2001–2005, and 2006–2010, respectively (*P*<0.0001 for each comparison) (Figure 1B, Table S4). This finding confirms prior belief of higher HSCR incidence in Blacks than Whites,^13,27^ but contradicts earlier report of highest incidence in Asians in the U.S.^26^ It also provides more precise population-level estimates based on significantly larger sample sizes in unbiased, national-wide data and highlights a significant health disparity in HSCR.

**Figure 1:**
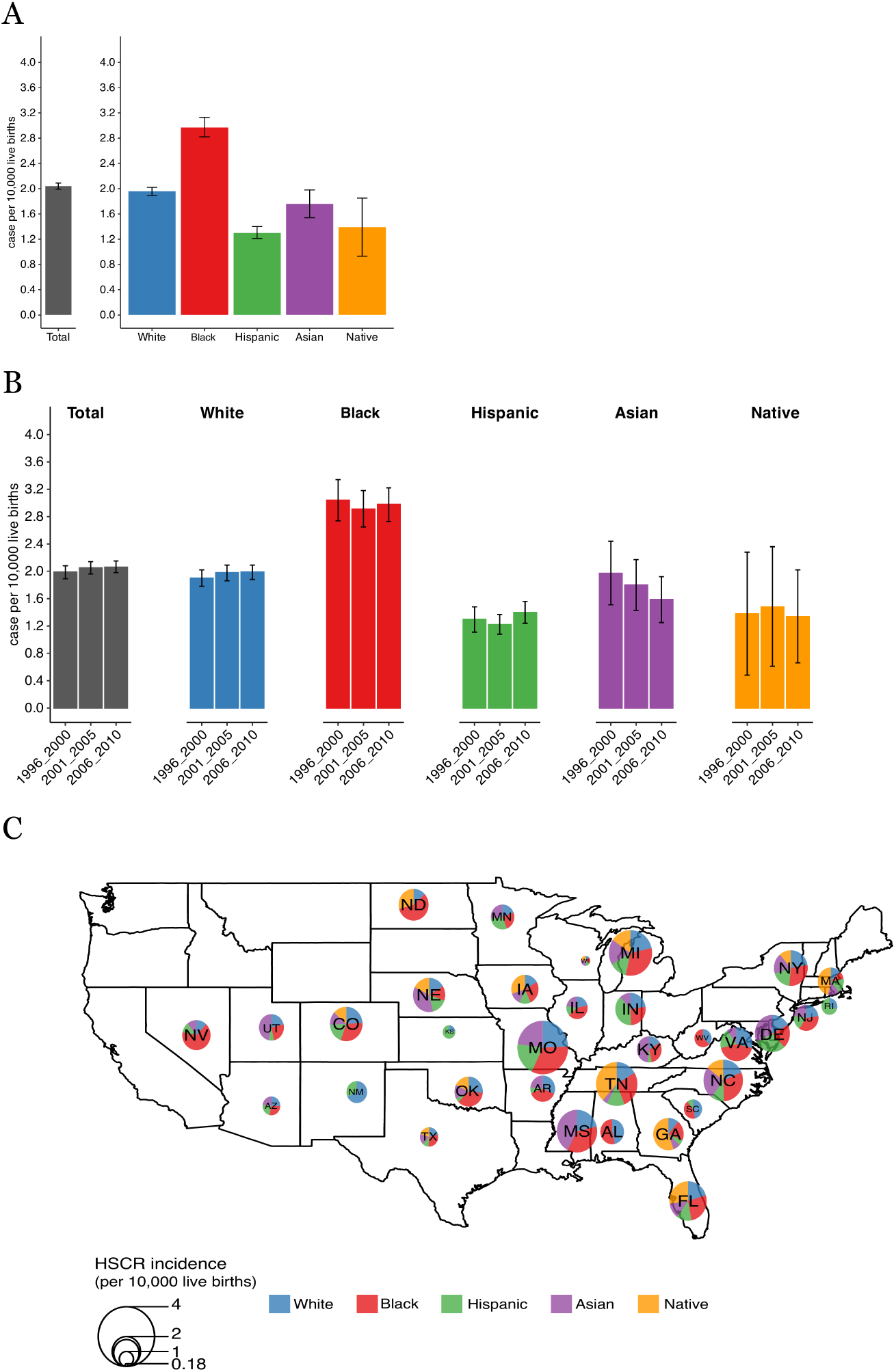
Population-specific incidence of Hirschsprung disease (HSCR): Data from the National Birth Defects Prevention Network (NBDPN; 1996–2010) (A) HSCR incidence in the U.S. based on the National Birth Defects Prevention Networks (NBDPN) data for the years 1996-2010. Information was extracted from 35 state-level surveillance data covering 30,236,567 individuals. (B) Total and population-specific HSCR incidence based on the NBDPN data, stratified by multiple surveillance time periods. (C) US geographic distribution of HSCR incidence by population based on the NBDPN data. Data from states with <1,000 live births at any surveillance interval were excluded to minimize outlier effects

HSCR incidence rates across SIRE groups were also compared across the U.S. geographically. Once again, the increased incidence in Blacks compared to Whites was consistent across states with 47.2% of the states reporting the highest incidence among Blacks. In contrast, 5.6%, 13.9%, 16.7%, and 16.7% of states showed the highest incidence among Whites, Hispanics, Asians, and Natives, respectively (Figure 1C). Multivariable regression analysis incorporating SIRE, temporal and geographic factors persistently identified SIRE as the main independent parameter associated with HSCR incidence variation, with Blacks being significantly associated with an increased HSCR incidence (β = 0.9, P= 2.1x10^-5^) after adjusting for time and location (Table S5).

To assess the generalizability of the incidence differences, we analyzed another independent nationwide data repository called Epic COSMOS. Incidence estimates from Epic COSMOS were concordant with those observed in NBDPN, including comparable overall HSCR incidence of 2.04 and 1.98 in NBDPN and Epic COSMOS, respectively, and similar SIRE-specific incidence patterns among Blacks and Whites (Black incidence = 2.98 and 2.80; White incidence =1.95 and 2.01, in NBDPN and Epic COSMOS, respectively) (Figure S3A, S3B).

For context, we further compared U.S. HSCR incidence with those reported in the literature. Overall HSCR incidence in the U.S. was comparable to global estimates (incidence _US_ = 1.99 – 2.09, incidence _global_ = 1.99 – 2.07, P= 0.76). Group-specific incidence was comparable for Whites (incidence _US_ = 1.89 – 2.02, incidence _global_ = 1.80 – 1.94, P= 0.11) but slightly lower for Asians (incidence _US_ = 1.54 – 1.98, incidence _global_ = 1.96 – 2.06, P= 0.03) (Figure S3C). Global incidences for Blacks and Hispanics were from a single U.S. study and thus not compared.^27^ This Asian incidence difference likely reflects population composition difference across study regions (Table S2, S3, S6), as East Asians are not identical to Asians in the U.S.

### Population Differences in HSCR Clinical Presentation

Clinical phenotype comparisons were then conducted using the HDRC cohort, with a final set of 237 White and 37 Black probands, in proportions consistent with U.S. census data (see Materials and Methods).

We compared multiple clinical features of HSCR, including diagnostic methods, mortality, demographic characteristics, clinical manifestations at HSCR diagnosis, HSCR-specific disease manifestations (e.g., segment length, familiality, and HSCR-associated syndromes, particularly Down syndrome), HSCR severity scores and overall complications, using their medical records. No group differences were identified in terms of diagnostic method, mortality, age at diagnosis, severity, sex ratio, familiality, segment length proportions, or HSCR-associated syndromes (Figure S4). However, we did identify differences in both initial clinical symptom manifestation at HSCR diagnosis and overall medical complications. At initial HSCR diagnosis, Whites manifested significantly more index symptoms: 56% with multiple index symptoms versus 35% in Blacks, respectively (P= 0.02, Figure 2A). Indeed, Black cases had a greater frequency of milder cases with chronic severe constipation as the sole manifestation with 16% in Blacks and 5% in Whites (P= 0.03, Figure 2B). Furthermore, White patients were more frequently (59%) reported to have other medical complications in addition to HSCR as compared to Blacks (40%) (P= 0.048, Figure 2C).

**Figure 2:**
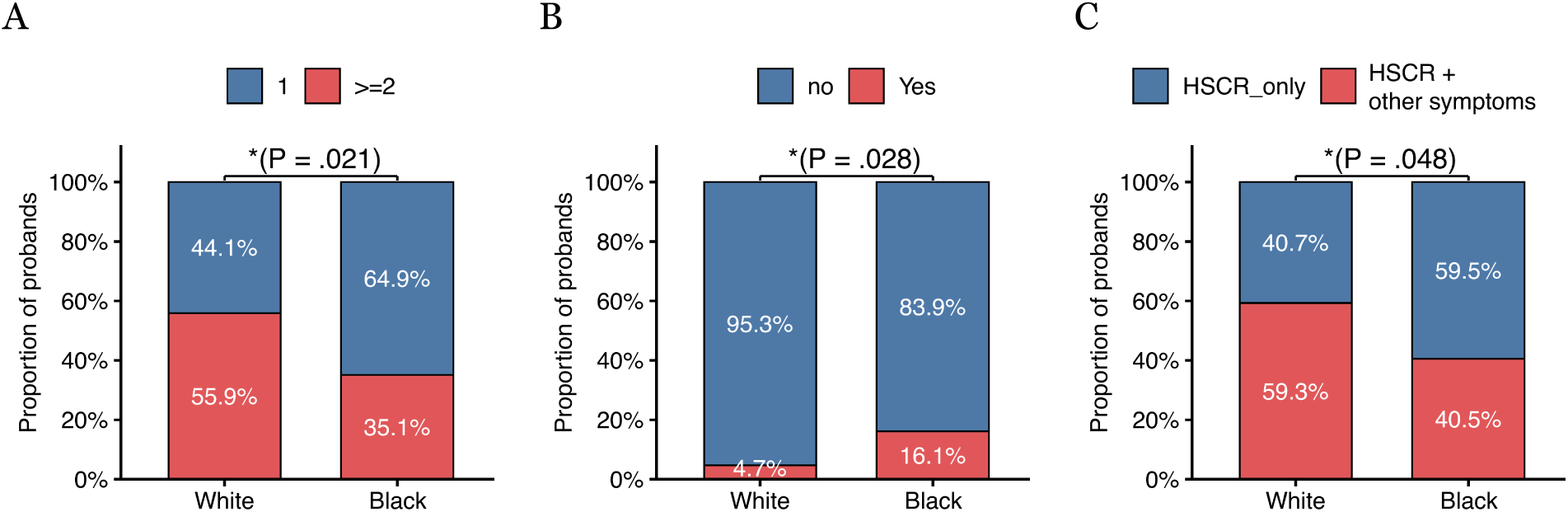
Comparisons of HSCR phenotypes and clinical presentations in non-Hispanic Blacks and Whites from the Hirschsprung Disease Research Consortium (HDRC) cohort. (A) Comparison of the fraction of major clinical symptoms used for HSCR diagnosis in 37 non-Hispanic Black and 237 non-Hispanic Whites. (B) Comparison of the proportion of cases with chronic severe constipation as the only index symptom for HSCR diagnosis, for the cases ascertained in (A). (C) Comparison of the fraction of HSCR cases with and without additional medical complications, for the cases ascertained in (A).

Taken together, these findings indicate that, within this nationally distributed cohort, Black patients more frequently presented with fewer and milder clinical manifestations, despite comparable diagnostic practices, demographic characteristics, and HSCR- specific disease features across these groups.

### SIRE Differences in Genetic Architecture

We next sought to ask whether any genetic differences in known HSCR genes and their regulatory elements could explain the milder presentation of HSCR in Blacks, using our existing whole genome sequence data. We focused our analyses on rare pathogenic coding variants (PVs) at 24 HSCR susceptibility genes and their transcriptional enhancer variants, given that their effects on HSCR are experimentally proven. ^38,80^

We first compared the cumulative burden of rare PVs on the two major genes for HSCR that explain the majority most of the disease’s susceptibility, namely, *RET* and *EDNRB*. Rare PVs were identified in 60 probands (54 White and 6 Black), comprising 53 distinct variants across these two genes. However, we did not detect any significant differences overall (P=0.75), nor when the data were stratified by variant class (loss-of-function P= 0.39, missense P= 0.47). Individually, neither *RET* nor *EDNRB* show any SIRE differences (P >0.05, Table S7).

Second, we compared the frequency profiles of the 10 experimentally validated enhancer variants at the *RET* locus and their haplotypes.^80^ In the HDRC cohort, all 10 enhancer variants were significantly associated with HSCR in White probands, whereas only 5 of them were significantly associated in Black probands (Table 1). Among the three major risk enhancer variants associated with known transcription factor binding – rs2506030 (*RET*−7), rs7069590 (*RET*−5.5), and rs2435357 (*RET*+3) – all were strongly associated in White probands (5.84 × 10^−31^, 6.84 × 10^−08^, 8.01 × 10^−62^, respectively), whereas only two (*RET*−7 and *RET*+3) reached statistical significance in Black probands (9.89 × 10^−06^, 2.88 × 10^−07^, respectively). Consistent with previous studies,^48,80^ the *RET*+3 (rs2435357) was the strongest risk variant with highest significance in both groups (Table 1). Note that given the sample size differences between the two samples of probands we did not expect identical levels of statistical significance.

**Table 1:** Population-specific case-control comparisons of 10 HSCR risk variants at the *RET* locus.

| SNP ID | Risk allele | Black/African American |  |  | White |  |  |
| --- | --- | --- | --- | --- | --- | --- | --- |
|  |  | Risk allele frequency: case | Risk allele frequency: control | P | Risk allele frequency: case | Risk allele frequency: control | P |
| rs788263 | G | 0.56 | 0.19 | **$2.41 \times 10^{-06}$** | 0.71 | 0.39 | **$8.46 \times 10^{-32}$** |
| rs788261 | C | 0.56 | 0.19 | **$2.44 \times 10^{-06}$** | 0.71 | 0.39 | **$8.55 \times 10^{-32}$** |
| rs788260 | A | 0.56 | 0.19 | **$2.42 \times 10^{-06}$** | 0.71 | 0.39 | **$8.14 \times 10^{-32}$** |
| rs2506030 | G | 0.54 | 0.16 | **$9.89 \times 10^{-06}$** | 0.71 | 0.39 | **$5.84 \times 10^{-31}$** |
| rs1547930 | G | 0.62 | 0.71 | $1.35 \times 10^{-01}$ | 0.86 | 0.75 | **$6.84 \times 10^{-08}$** |
| rs7069590 | T | 0.64 | 0.56 | $2.17 \times 10^{-01}$ | 0.84 | 0.76 | **$1.94 \times 10^{-04}$** |
| rs2435357 | T | 0.23 | 0.05 | **$2.88 \times 10^{-07}$** | 0.65 | 0.25 | **$8.01 \times 10^{-62}$** |
| rs12247456 | G | 0.58 | 0.45 | $4.71 \times 10^{-02}$ | 0.81 | 0.67 | **$1.73 \times 10^{-09}$** |
| rs7393733 | G | 0.58 | 0.45 | $4.70 \times 10^{-02}$ | 0.81 | 0.67 | **$2.26 \times 10^{-09}$** |
| rs2505541 | T | 0.79 | 0.70 | $1.76 \times 10^{-01}$ | 0.83 | 0.59 | **$1.33 \times 10^{-26}$** |

The table shows the frequency of risk alleles in cases and controls for each population. P values were calculated using Fisher's exact test; statistically significant values are bolded. The cases were from the HDRC cohort and controls were from gnomADv4 (i.e., gnomAD Black/African American and gnomAD non-Finnish European).

Haplotype analyses also demonstrated differences in *RET* enhancer variant burden by SIRE. In Whites, multiple haplotypes showed strong association with HSCR (CTGA**GTTGGT**: OR=3.3, 95%CI: 1.86–5.87, P= 3.38 × 10^−05^ and **GCAGGTTGGT**: OR=4.91, 95%CI: 2.94–8.20, P= 7.48 × 10^−12^, risk alleles are bolded), whereas only one was nominally significant in Blacks but not after Bonferroni correction (**GCAGGTTGGT**: OR=5.4, 95%CI: 1.18–24.65, P=0.04) (Table 2). In contrast, when restricted to the 5 enhancer variants significant in both groups of probands, two haplotypes were associated with HSCR in both groups, although the significance were consistently larger in Whites (CTGA**T**: OR=6.34, 95%CI: 1.63–24.63, P= 1.06 × 10^−02^ in Blacks and OR=5.05, 95%CI: 3.59–7.1, P=1.64 × 10^−19^in Whites; **GCAGT**: OR=5.07, 95%CI: 1.99–12.97, P= 1.07 × 10^−03^ in Blacks and OR=6.94, 95%CI: 5.22–9.23, P=2.84 × 10^−43^in Whites) (Table S8). Thus, considering the sample size difference, regulatory variant effects in *RET* are not different between Blacks and Whites.

**Table 2:** Comparisons of 10 risk enhancer variant haplotypes at the *RET* locus by population.

| Haplotype | # Risk alleles | Black/African American |  |  |  | White |  |  |  |
| --- | --- | --- | --- | --- | --- | --- | --- | --- | --- |
|  |  | case<br>freq | ctrl<br>freq | OR<br>(95% CI) | P | case<br>freq | ctrl<br>freq | OR<br>(95% CI) | P |
| CTGAACCACT | 1 | 0.05 | 0.07 | (1=ref) | — | 0.05 | 0.07 | (1=ref) | — |
| CTGAACCGGC | 2 | 0.03 | 0.03 | 1.50 | 1.00 | 0.01 | 0.01 | 2.07 | 0.39 |
| CTGAATCACT | 2 | 0.06 | 0.07 | 1.26 | 1.00 | 0.03 | 0.08 | 0.61 | 0.26 |
| CTGAGCCACT | 2 | 0.14 | 0.19 | 1.08 | 1.00 | 0.05 | 0.09 | 0.89 | 0.74 |
| CTGAATCGGC | 3 | 0.09 | 0.05 | 3.00 | 0.26 | 0.01 | 0.06 | **0.35**<br>**(0.13–0.97)** | **0.049** |
| CTGAGTCACT | 3 | 0.08 | 0.08 | 1.43 | 0.72 | 0.01 | 0.01 | 0.78 | 1.00 |
| CTGAGTCGGC | 4 | 0.06 | 0.16 | 0.59 | 0.67 | 0.05 | 0.15 | 0.53 | 0.079 |
| CTGAGTTGGT | 6 | 0.03 | 0.01 | 6.00 | 0.17 | 0.13 | 0.06 | **3.3**<br>**(1.86–5.87)** | **3.4×10<sup>-5</sup>** |
| GCAGGCCACT | 6 | 0.02 | 0.03 | 0.75 | 1.00 | 0.02 | 0.04 | 0.80 | 0.68 |
| GCAGGTCACT | 7 | 0.02 | 0.03 | 0.75 | 1.00 | 0.03 | 0.04 | 1.17 | 0.69 |
| GCAGGTTGGT | 10 | 0.14 | 0.04 | **5.4**<br>**(1.18–24.65)** | **0.038** | 0.44 | 0.14 | **4.91**<br>**(2.94–8.20)** | **7.5×10<sup>-12</sup>** |
| CTGAACCGGT | 3 | 0.03 | 0.06 | 0.75 | 1.00 | — | — | — | — |
| CTGAATCGGT | 4 | 0.03 | 0.02 | 2.40 | 0.57 | — | — | — | — |
| CTGAGCCGGT | 4 | 0.05 | 0.02 | 4.50 | 0.14 | — | — | — | — |
| GCAAGCCACT | 5 | 0.02 | 0.01 | 2.00 | 0.53 | — | — | — | — |
| CTGAGTCGGT | 5 | 0.02 | 0.03 | 0.86 | 1.00 | — | — | — | — |
| GCAAGTCGGC | 7 | 0.03 | 0.01 | 6.00 | 0.17 | — | — | — | — |
| CTGAGCCGGC | 3 | — | — | — | — | 0.02 | 0.04 | 0.74 | 0.66 |
| CTGAATTGGT | 5 | — | — | — | — | 0.04 | 0.02 | **3.16**<br>**(1.46–6.81)** | **4.2×10<sup>-3</sup>** |
| GCAGGTCGGC | 8 | — | — | — | — | 0.07 | 0.15 | 0.75 | 0.42 |

Observed frequencies of haplotypes and counts (N) in cases and controls, together with the odds ratio (with respect to the reference haplotype containing the fewest risk alleles), are shown for 33 Black/African American cases, 204 White cases, 129 Black/African American controls and 970 White controls. Risk alleles are bolded. P values were calculated using Fisher's exact test; significant values are bolded. The 10 risk enhancer variants in each haplotype are in order: rs788263 |rs788261 |rs788260 |rs2506030 |rs1547930 |rs7069590 |rs2435357 |rs12247456 |rs7393733 |rs25055

### Group difference in putative enhancer variants at *SOX10*

The small, quantitative and non-significant enhancer effect size differences observed above prompted us to explore whether enhancers of the other 23 HSCR genes could show differences. Of these, only *SOX10* demonstrated strong enrichment of common variants in Black probands as compared to Whites (Figure 3A; Supplementary Table S9).

**Figure 3:**
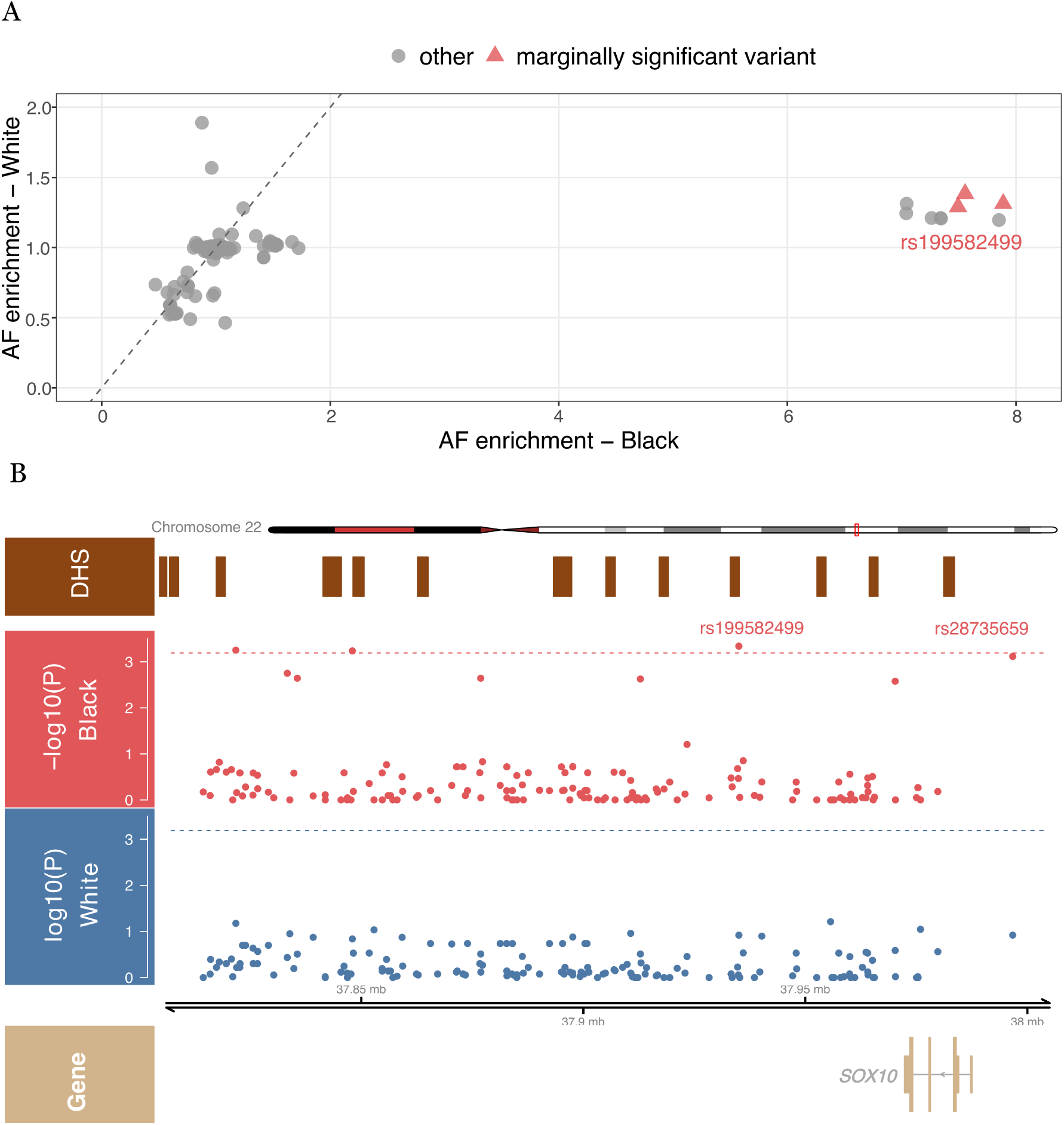
Common non-coding variants at the *SOX10* locus by population, overlaid on DNase I hypersensitive sites (DHS) from human embryonic gut tissues. (A) Enrichment of allele frequency (AF) for each variant in Black and White cases as compared to controls. The latter are from the gnomAD database with enrichment calculated as a case- control ratio. (B) The specific regulatory elements (DHS), the variant level statistical association (-log10(P value)) for case-control allele frequency comparison in Blacks and Whites, and the genomic location of *SOX10* canonical transcript are indicated. Marginally significant levels are highlighted with doted lines.

In Black probands, three less common variants at *SOX10* reached marginal significance, whereas no variants were enriched in White probands (Figure 3B). One putative enhancer variant, rs199582499, showed an 8-fold allele enrichment in Black cases as compared with controls (cases frequency = 8.1%; control frequency = 1.0%; *P=*4.58×10⁻⁴) but it was not significantly enriched in Whites (cases frequency 8.1%; control frequency 6.2%; *P=*0.12) despite the larger allele frequencies and sample size (Figure 3A). This variant lies within a putative regulatory functional region based on chromatin accessibility (DHS) data from human embryonic colon tissue and is located 35.6 kb downstream of *SOX10*. Additional analyses identified a second putative enhancer variant, rs28735659, located 9.3 kb upstream of *SOX10*, which lies within an experimentally validated functional CRE of *SOX10*.^86^ This variant is also significantly enriched in Black (case frequency = 7.6%; control frequency = 1.1%; *P=*7.66×10⁻⁴) but not in White (case frequency = 8.1%, control frequency = 6.2%, P= 0.12) cases (Figure 3A). Indeed, the two putative *SOX10* enhancer variants are in high allelic association with one another in both African- (R^2^ =1; ASW) and European-ancestry (R^2^ =0.8; CEU) subjects based on the 1000 Genomes reference populations. One may speculate whether this regulatory variant is one cause of the milder presentation in Blacks as well as a reason for its higher incidence.

## Discussion

In this study, we used multiple data sources, to address US population, particularly Black-White, differences, using population-based birth defects surveillance data, and a repository of electronic health records data. We demonstrate significant differences in the incidence of HSCR. We found that the overall incidence of HSCR is 2.04 per 10,000 live births, consistent with the usually assumed 1/5,000 live births incidence used by most authors for European-ancestry and U.S. White subjects.^8^ However, our data definitively show that Blacks have the highest HSCR incidence among U.S. population groups, consistent with a recent study from a California-based birth defects registry, which was not part of the NBDPN.^27^ Analogously, Asians in the U.S. have the lowest incidence, lower than Whites, although not statistically significantly so, while rates in Hispanics and Native Americans are lower still. We believe these patterns are real and not artefactual because they are consistent across birth cohorts over time, across U.S. states and geographic regions, and consistent across both nationwide datasets, despite the datasets not spanning identical time periods. The causes of these population differences are, however, unknown. HSCR is a multifactorial disease with both genetic and suspected environmental risk factors. To date, the genetic risk factors have major effects, are well recognized and their specifics have been elucidated by many studies. ^38,39^ Environmental risk factors are indeed plausible but have not been definitively proven.^33^ Nevertheless, population differences may arise from either genes or the environment or both. Importantly, the consistently higher HSCR incidence among Blacks observed across U.S. states and regions points toward broad social and structural determinants and genetics, rather than localized environment factors. However, the 50% incidence increase in U.S. Blacks relative to Whites is a major difference and may even be under- estimated due to healthcare access^87^ and research study ascertainment differences.^88,89^

Intriguingly, despite this higher incidence, Black patients more frequently present with milder clinical features of HSCR, such as higher rates of its milder form of chronic severe constipation. They are also more likely to have HSCR as the sole clinical condition reported in their EHR. At the genetic level, no significant differences were observed in the burden of rare pathogenic coding variants between Black and White patients. Instead, small genetic differences were primarily observed in the HSCR risk gene noncoding regulatory architecture, with group-specific patterns at *RET* and *SOX10*, although the clinical consequences of these differences remain unclear. In addition, we identified two putative enhancer variants at *SOX10* unique to Black patients. Pathogenic coding variants at *SOX10* leads to syndromic HSCR.^8,40^ The observed noncoding variants are expected to decrease *SOX10* gene expression, thereby leading to decreased *SOX10* protein expression and lower *RET* gene expression.^39,80^ The association of these *SOX10* enhancer variants with milder disease in Blacks suggests that perturbing *SOX10* through its enhancers may have a smaller effect on *RET* gene expression than variants in the *SOX10* binding enhancer of *RET*.^80,90^ Whether similar regulatory variants exist in other populations remains unclear as HSCR cases have been limitedly sampled in other groups. Together, these findings emphasize how the study of diverse populations beyond addressing health disparities can illuminate genotype-phenotype relationships.

Beyond genetic explanations, these data also highlight the need for considering population differences in clinical practice and phenotyping. Thus, it’s feasible that Black patients have not been clinically examined as thoroughly as White patients. We need to conduct specific studies to quantify the degree of clinical attention provided to diverse patients and the impact of them on which patients get ascertained for research studies.

Population and SIRE differences in clinical presentations are not unique to HSCR. Indeed, such phenotype heterogeneity has been reported in diverse pediatric and rare diseases, particularly for complex or multifactorial disorders.^91–93^ Such differences are generally attributed to the combined effects of social, environmental and genetic factors. Social and structural determinants of health may influence the clinical outcomes through underlying health conditions, environmental exposures and disease vulnerability across populations.^93–95^ However, the phenotypic heterogeneity observed in this study likely stems from population-specific differences in the genomic landscape as well.

Genetic features such as variation in risk allele and haplotype frequencies can lead to different genetic backgrounds that modify the disease penetrance and severity differentially.^96–98^ In HSCR, where the genetic architecture is multi-factorial and has been demonstrated to involve synergistic effects of both rare and common variants,^38,39,99^ these background genetic differences can lead to quantitatively differential risk across populations. These differences are more likely to arise from common disease variants, in theory, as opposed to rare disease variants, because the frequency of the latter are almost entirely determined by genetic parameters (gene mutation rate and selection) that are generally independent of population origin.^100^ On the other hand, the former is largely determined by historical population demography that can vary significantly between groups.^101^ The fact that the large majority of complex disease risk arises from regulatory variation, in HSCR and elsewhere, is consistent with this model. This aligns with a prior study indicating that a substantial portion of individual genetic variations is from the difference in transcription factor (TF) binding.^102^ Indeed, we already know that the many enhancer variants in HSCR that contribute to risk disrupt known TFs and show wide population variation in their frequencies.^39,49,68,80^

This study has several strengths. First, it integrates population-based epidemiologic surveillance, detailed clinical phenotyping, and genetic analyses within a unified analytic framework, enabling direct comparison of SIRE differences in HSCR incidence, clinical presentation, and genetic architecture. Second, the use of independent, large, multisite, nationwide data sources minimizes the likelihood that our findings are driven by single-center or regional effects. Finally, the availability of in-depth medical records alongside genome sequencing data from a cohort of patients enables exploration of how genetic variation, particularly noncoding regulatory variants may relate to disease clinical manifestations.

This study also has some limitations. Although the HDRC cohort represents one of the largest and most diverse HSCR collections in the U.S., the number of Black and other non-majority patients remains modest, which limits the statistical power to detect genetic and environmental differences in them. In particular, the smaller numbers of Asian, Hispanic, and Native American cases in our studies precluded adequately powered subgroup-specific genetic analyses. While our cohort has a White-Black composition comparable to that of the U.S. population, residual ascertainment bias cannot be entirely excluded. More broadly, the under-representation of minority populations in rare disease research remains a major challenge,^93–95^ and limited sample sizes reduce power and increase susceptibility to random confounding. Finally, these analyses are focused on U.S.-based populations, and the findings may not be generalizable to non-U.S. populations in other regions or healthcare settings.

This study has several implications for future research and clinical understanding of HSCR. First, it highlights the importance of SIRE- or genetic-ancestry stratified, population-aware analyses for HSCR and other multifactorial pediatric rare diseases, as aggregate estimates obscures meaningful heterogeneity in disease burden, clinical presentation, and genetic architecture. The identification of population-specific noncoding regulatory variants, particularly the ones at *SOX10*, also underscores the need to expand genetic studies beyond coding regions and beyond European-ancestry and White populations. Current HSCR genetic testing largely focuses on coding variants, and our results suggest that common noncoding enhancer variants have greater contributions to both disease risk and severity and should be evaluated. Future studies using larger, more diverse cohorts, along with functional validations for genetic discoveries will be essential to clarify the connections among disease risk, clinical presentation and genetics. In the meantime, this study has shown that closer scrutiny of African-ancestry patients is warranted.

## Data Availability

All data produced in the present study are available upon reasonable request to the authors

## Data Availability

Access to individual-level data is subject to institutional approvals and applicable data-use agreements. Aggregate results and supplementary materials are provided in the Supplement. Requests for additional data may be available through Kids First Data Resource Portal with dbGaP application or directed to Dr. Aravinda Chakravarti, and may require additional IRB permissions and therefore take time.

## Author Contributions

M.F. and A.C. conceptualized and designed the study. M.F., H.B.-R and M.Z prepared the samples for sequencing. M.F. performed the analyses and drafted the manuscript. S.C. and H.B.-R. contributed to the study design and analysis pipelines. All authors reviewed the results, provided critical feedback, and contributed to the final manuscript.

## Acknowledgement

We thank all the patients and families who participated in HDRC study. We acknowledge the Gabriella Miller Kids First Pediatric Research Program (project HD110884-01) for their sequencing capabilities.

## References

1. Bolande, R. The neurocristopathies A unifying concept of disease arising in neural crest maldevelopment. Hum. Pathol. 5, 409–429 (1974).

2. Taraviras, S. & Pachnis, V. Development of the mammalian enteric nervous system. Curr. Opin. Genet. Dev. 9, 321–327 (1999).

3. Whitehouse, F. R. & Kernohan, J. W. Myenteric plexus in congenital megacolon; study of 11 cases. Arch. Intern. Med. (Chic). 82, 75–111 (1948).

4. Ambartsumyan, L., Smith, C. & Kapur, R. P. Diagnosis of Hirschsprung Disease. Pediatric and Developmental Pathology 23, 8–22 (2020).

5. Amiel, J. Hirschsprung disease, associated syndromes, and genetics: a review. J. Med. Genet. 38, 729–739 (2001).

6. Badner, J. A., Sieber, W. K., Garver, K. L. & Chakravarti, A. A genetic study of Hirschsprung disease. Am. J. Hum. Genet. 46, 568–580 (1990).

7. Kyrklund, K. et al. ERNICA guidelines for the management of rectosigmoid Hirschsprung’s disease. Orphanet J. Rare Dis. 15, 164 (2020).

8. *Chakravarti, A., McCallion, A. S. & Lyonnet, S. Hirschsprung Disease. The Online Metabolic & Molecular Bases of Inherited Disease DOI: 10.1036/ommbid.291 (2019) doi:DOI: 10.1036/ommbid.291.

9. Hartman EE et al. Critical factors affecting quality of life of adult patients with anorectal malformations or Hirschsprung’s disease. Am J Gastroenterol 99: 907–913, 2004.

10. Hartman EE et al. Explaining change over time in quality of life of adult patients with anorectal malformations or Hirschsprung’s disease. Dis Colon Rectum 49: 96–103, 2006.

11. Menezes M, Corbally M, Puri P. Long-term results of bowel function after treatment for Hirschsprung’s disease: a 29-year review. Pediatr Surg Int 22:987–90, 2006.

12. Le-Nguyen, A., Righini-Grunder, F., Piché, N., Faure, C. & Aspirot, A. Factors influencing the incidence of Hirschsprung associated enterocolitis (HAEC). J. Pediatr. Surg. 54, 959–963 (2019).

13. Goldberg, E. L. An Epidemiological Study of Hirschsprung ’s Disease. 13, (1984).

14. Best, K. E., Glinianaia, S. V., Bythell, M. & Rankin, J. Hirschsprung’s disease in the North of England: Prevalence, associated anomalies, and survival. Birth Defects Res. A Clin. Mol. Teratol. 94, 477–480 (2012).

15. Best, K. E. et al. Hirschsprung’s disease prevalence in Europe: A register-based study. Birth Defects Res. A Clin. Mol. Teratol. 100, 695–702 (2014).

16. Bradnock, T. J., Knight, M., Kenny, S., Nair, M. & Walker, G. M. Hirschsprung’s disease in the UK and Ireland: Incidence and anomalies. Arch. Dis. Child. 102, 722–727 (2017).

17. Downey, E. C., Hughes, E., Putnam, A. R., Baskin, H. J. & Rollins, M. D. Hirschsprung disease in the premature newborn: A population based study and 40-year single center experience. J. Pediatr. Surg. 50, 123–125 (2015).

18. Nasr, A., Sullivan, K. J., Chan, E. W., Wong, C. A. & Benchimol, E. I. Validation of algorithms to determine incidence of hirschsprung disease in ontario, Canada: A population-based study using health administrative data. Clin. Epidemiol. 9, 579–590 (2017).

19. Russell, M. B., Russell, C. A. & Niebuhr, E. An epidemiological study of Hirschsprung’s disease and additional anomalies. Acta Paediatrica, International Journal of Paediatrics 83, 68–71 (1994).

20. Spouge, D. & Baird, P. A. Hirschsprung disease in a large birth cohort. Teratology 32, 171–177 (1985).

21. Chia, S. T., Chen, S. C. C., Lu, C. L., Sheu, S. M. & Kuo, H. C. Epidemiology of Hirschsprung’s Disease in Taiwanese Children: A 13-year Nationwide Population-based Study. Pediatr. Neonatol. 57, 201–206 (2016).

22. Ikeda, K. & Goto, S. Diagnosis and Treatment of Hirschsprung’s Disease in Japan. Ann. Surg. 199, 400–405 (1984).

23. Suita, S., Taguchi, T., Ieiri, S. & Nakatsuji, T. Hirschsprung’s disease in Japan: analysis of 3852 patients based on a nationwide survey in 30 years. J. Pediatr. Surg. 40, 197–202 (2005).

24. Taguchi, T., Obata, S. & Ieiri, S. Current status of Hirschsprung’s disease: based on a nationwide survey of Japan. Pediatr. Surg. Int. 33, 497–504 (2017).

25. Rajab, A., Freeman, N. V. & Patton, M. A. Hirschsprung’s disease in Oman. J. Pediatr. Surg. 32, 724–727 (1997).

26. Torfs, C. P. An epidemiological study of Hirschsprung’disease in a multiracial California population. Third International Meeting: Hirschsprung’s disease and related neurocristophaties Evian, France (1998).

27. Anderson, J. E. et al. Epidemiology of Hirschsprung disease in California from 1995 to 2013. Pediatr. Surg. Int. 34, 1299–1303 (2018).

28. Lipson, A. Hirschsprung disease in the offspring of mothers exposed to hyperthermia during pregnancy. Am. J. Med. Genet. 29, 117–24 (1988).

29. Larsson, L. T., Okmian, L. & Kristoffersson, U. No correlation between hyperthermia during pregnancy and Hirschsprung disease in the offspring. Am. J. Med. Genet. 32, 260–1 (1989).

30. Fu, M. et al. Vitamin A facilitates enteric nervous system precursor migration by reducing Pten accumulation. Development 137, 631–40 (2010).

31. Schill, E. M. et al. Ibuprofen slows migration and inhibits bowel colonization by enteric nervous system precursors in zebrafish, chick and mouse. Dev. Biol. 409, 473–88 (2016).

32. Lake, J. I., Tusheva, O. A., Graham, B. L. & Heuckeroth, R. O. Hirschsprung-like disease is exacerbated by reduced de novo GMP synthesis. J. Clin. Invest. 123, 4875–87 (2013).

33. Heuckeroth, R. O. & Schäfer, K.-H. Gene-environment interactions and the enteric nervous system: Neural plasticity and Hirschsprung disease prevention. Dev. Biol. 417, 188–97 (2016).

34. Beltman, L. et al. Diagnosing Hirschsprung Disease in Children Younger than 6 Months of Age: Insights in Incidence of Complications of Rectal Suction Biopsy and Other Final Diagnoses. (2023).

35. Qualman, S. J., Jaffe, R., Bove, K. E. & Monforte-Muñoz, H. Diagnosis of Hirschsprung disease using the rectal biopsy: Multi-institutional survey. Pediatric and Developmental Pathology 2, 588–596 (1999).

36. Ho, J. M. D. & How, C. H. Chronic constipation in infants and children. Singapore Med. J. 61, 63–68 (2020).

37. Hyams, J. S. et al. Childhood Functional Gastrointestinal Disorders: Child/Adolescent. Gastroenterology 150, 1456–1468.e2 (2016).

38. Tilghman, J. M. et al. Molecular Genetic Anatomy and Risk Profile of Hirschsprung’s Disease. N. Engl. J. Med. 380, 1421–1432 (2019).

39. Chatterjee, S., et al. Enhancer Variants Synergistically Drive Dysfunction of a Gene Regulatory Network In Hirschsprung Disease. Cell 167, 355–368.e10 (2016).

40. Pingault, V. et al. SOX10 mutations in patients with Waardenburg-Hirschsprung disease. Nat. Genet. 18, 171–3 (1998).

41. Luzón-Toro, B. et al. Exome sequencing reveals a high genetic heterogeneity on familial Hirschsprung disease. Sci. Rep. 5, 16473 (2015).

42. Fu, A. X. et al. Whole-genome analysis of noncoding genetic variations identifies multiscale regulatory element perturbations associated with Hirschsprung disease. Genome Res. 30, 1618–1632 (2020).

43. Tang, C. S.-M. et al. Identification of Genes Associated With Hirschsprung Disease, Based on Whole-Genome Sequence Analysis, and Potential Effects on Enteric Nervous System Development. Gastroenterology 155, 1908–1922.e5 (2018).

44. Gui, H. et al. Whole exome sequencing coupled with unbiased functional analysis reveals new Hirschsprung disease genes. Genome Biol. 18, 48 (2017).

45. Angrist, M. et al. Mutation analysis of the RET receptor tyrosine kinase in Hirschsprung disease. Hum. Mol. Genet. 4, 821–830 (1995).

46. Attié, T. et al. Diversity of RET proto-oncogene mutations in familial and sporadic Hirschsprung disease. Hum. Mol. Genet. 4, 1381–6 (1995).

47. Garcia-Barceló, M., et al. Highly Recurrent RET Mutations and Novel Mutations in Genes of the Receptor Tyrosine Kinase and Endothelin Receptor B Pathways in Chinese Patients with Sporadic Hirschsprung Disease. Clin. Chem. 50, 93–100 (2004).

48. Emison, E. S. et al. A common sex-dependent mutation in a RET enhancer underlies Hirschsprung disease risk. Nature 434, 857–863 (2005).

49. Kapoor, A. et al. Population variation in total genetic risk of Hirschsprung disease from common RET, SEMA3 and NRG1 susceptibility polymorphisms. Hum. Mol. Genet. 24, 2997–3003 (2015).

50. Office of Management and Budget. Revisions to the Standards for the Classification of Federal Data on Race and Ethnicity. Fed. Regist. (1997).

51. Noel, A. & Bartelt, K. Cosmos: Real-World Data Powered by the Healthcare Community. Journal of the Society for Clinical Data Management 3, (2023).

52. By, E. & Sever, L. E. Guidelines for Conducting Birth Defects Surveillance NATIONAL BIRTH DEFECTS PREVENTION NETWORK. http://www.nbdpn.org/bdsurveillance.html. (2004).

53. Saini, V., Jaber, T., Como, J. D., Lejeune, K. & Bhanot, N. 623. Exploring ‘Slicer Dicer’, an Extraction Tool in EPIC, for Clinical and Epidemiological Analysis. Open Forum Infect. Dis. 8, S414–S415 (2021).

54. Stephens, F. D. The diagnosis and management of Hirschsprung’s disease. Ann. R. Coll. Surg. Engl. 7, 257–68 (1950).

55. Tafuto, B. & Lechner, D. W. REDCap as an accreditation tool for academic programs in clinical research: A case study. J. Clin. Transl. Sci. 8, e185 (2024).

56. Steinlechner, M. & Parson, W. Automation and high through-put for a DNA database laboratory: Development of a Laboratory Information Management System. Croat. Med. J. 42, 252–255 (2001).

57. Murphy, K. & Wolf, T. Tools of the Trade: CONFIDENCE INTERVALS for a CRUDE RATE. 1–3 (2015).

58. Minassian, D. & Kuper, H. Cross-sectional studies. The Epidemiology of Eye Disease 79–93 (2012) doi:10.1142/9781848166271_0003.

59. Altman, D. G. Practical Statistics for Medical Research. (Chapman and Hall/CRC, 1990). doi:10.1201/9780429258589.

60. Byrska-Bishop, M. et al. High-coverage whole-genome sequencing of the expanded 1000 Genomes Project cohort including 602 trios. Cell 185, 3426–3440.e19 (2022).

61. Bryc, K., Durand, E. Y., Macpherson, J. M., Reich, D. & Mountain, J. L. The genetic ancestry of african americans, latinos, and european Americans across the United States. Am. J. Hum. Genet. 96, 37–53 (2015).

62. Price, A. L. et al. Discerning the Ancestry of European Americans in Genetic Association Studies. PLoS Genet. 4, e236 (2008).

63. Lao, O. et al. Evaluating self-declared ancestry of U.S. Americans with autosomal, Y-chromosomal and mitochondrial DNA. Hum. Mutat. 31, E1875–E1893 (2010).

64. Bryc, K. et al. Genome-wide patterns of population structure and admixture in West Africans and African Americans. Proc. Natl. Acad. Sci. U. S. A. 107, 786–791 (2010).

65. Tishkoff, S. A., et al. The Genetic Structure and History of Africans and African Americans. Science (1979). 324, 1035–1044 (2009).

66. Zakharia, F. et al. Characterizing the admixed African ancestry of African Americans. Genome Biol. 10, (2009).

67. Eric Jensen, Nicholas Jones, Kimberly Orozco, Lauren Medina, M. P. Measuring Racial and Ethnic Diversity for the 2020 Census. Census.gov https://www.census.gov/newsroom/blogs/randomsamplings/2021/08/measuring-racial-ethnic-diversity-2020-census.html (2022).

68. Kapoor, A. et al. Multiple, independent, common variants at RET, SEMA3 and NRG1 gut enhancers specify Hirschsprung disease risk in European ancestry subjects. J. Pediatr. Surg. 56, 2286–2294 (2021).

69. Hu, T., Chitnis, N., Monos, D. & Dinh, A. Next-generation sequencing technologies: An overview. Hum. Immunol. 82, 801–811 (2021).

70. GATK Team. Genotype Refinement workflow for germline short variants. https://gatk.broadinstitute.org/hc/en-us/articles/360035531432-Genotype-Refinement-workflow-for-germline-short-variants (2024).

71. Fu, M., Berk-Rauch, H. E., Chatterjee, S. & Chakravarti, A. The Role of de novo and Ultra-Rare Variants in Hirschsprung Disease (HSCR): Extended Gene Discovery for Risk Profiling of Patients. medRxiv 10.1101/2025.01.07.25320162 (2025) doi:10.1101/2025.01.07.25320162.

72. Gudmundsson, S. et al. Variant interpretation using population databases: Lessons from gnomAD. Hum. Mutat. 43, 1012–1030 (2022).

73. Rao, S. S. P. et al. A 3D Map of the Human Genome at Kilobase Resolution Reveals Principles of Chromatin Looping. Cell 162, 687–688 (2015).

74. Meuleman, W. et al. Index and biological spectrum of human DNase I hypersensitive sites. Nature 584, 244–251 (2020).

75. ENCODE Project Consortium. An integrated encyclopedia of DNA elements in the human genome. Nature 489, 57–74 (2012).

76. Faure, A. J. et al. Cohesin regulates tissue-specific expression by stabilizing highly occupied cis-regulatory modules. Genome Res. 22, 2163–2175 (2012).

77. Thurman, R. E. et al. The accessible chromatin landscape of the human genome. Nature 489, 75–82 (2012).

78. Hei HA, J. L., Hang LUI, V. C. & Hang TAM, P. K. Embryology and anatomy of Hirschsprung disease. Semin. Pediatr. Surg. 31, 151227 (2022).

79. Mudge, J. M., et al. GENCODE 2025: reference gene annotation for human and mouse. Nucleic Acids Res. 53, D966–D975 (2025).

80. Chatterjee, S., Karasaki, K. M., Fries, L. E., Kapoor, A. & Chakravarti, A. A multi-enhancer RET regulatory code is disrupted in Hirschsprung disease. Genome Res. 31, 2199–2208 (2021).

81. Browning, B. L., Tian, X., Zhou, Y. & Browning, S. R. Fast two-stage phasing of large-scale sequence data. The American Journal of Human Genetics 108, 1880–1890 (2021).

82. Hofmeister, R. J., Ribeiro, D. M., Rubinacci, S. & Delaneau, O. Accurate rare variant phasing of whole-genome and whole-exome sequencing data in the UK Biobank. Nat. Genet. 55, 1243–1249 (2023).

83. Lonsdale, J. et al. The Genotype-Tissue Expression (GTEx) project. Nat. Genet. 45, 580–585 (2013).

84. Barrett, J. C., Fry, B., Maller, J. & Daly, M. J. Haploview: analysis and visualization of LD and haplotype maps. Bioinformatics 21, 263–5 (2005).

85. Cheverud, J. M. A simple correction for multiple comparisons in interval mapping genome scans. Heredity (Edinb). 87, 52–8 (2001).

86. Antonellis, A. et al. Identification of Neural Crest and Glial Enhancers at the Mouse Sox10 Locus through Transgenesis in Zebrafish. PLoS Genet. 4, e1000174 (2008).

87. Yearby, R. Racial Disparities in Health Status and Access to Healthcare: The Continuation of Inequality in the United States Due to Structural Racism. The American Journal of Economics and Sociology 77, 1113–1152 (2018).

88. Martin, A. R. et al. Human Demographic History Impacts Genetic Risk Prediction across Diverse Populations. The American Journal of Human Genetics 100, 635– 649 (2017).

89. Martin, A. R. et al. Clinical use of current polygenic risk scores may exacerbate health disparities. Nat. Genet. 51, 584–591 (2019).

90. Emison, E. S. et al. Differential Contributions of Rare and Common, Coding and Noncoding Ret Mutations to Multifactorial Hirschsprung Disease Liability. The American Journal of Human Genetics 87, 60–74 (2010).

91. Onamusi, T., Murphy, J. & Shah, S. D. Racial Differences in Disease Characteristics of Pediatric Hidradenitis Suppurativa. Pediatr. Dermatol. 42, 806–809 (2025).

92. Weiler, T., Mikhail, I., Singal, A. & Sharma, H. Racial Differences in the Clinical Presentation of Pediatric Eosinophilic Esophagitis. J. Allergy Clin. Immunol. Pract. 2, 320–325 (2014).

93. Fiscella, K. & Sanders, M. R. Racial and Ethnic Disparities in the Quality of Health Care. Annu. Rev. Public Health 37, 375–394 (2016).

94. Briscoe, S. et al. Evidence of inequities experienced by the rare disease community with respect to receipt of a diagnosis and access to services: a scoping review of UK and international evidence. Orphanet J. Rare Dis. 20, 303 (2025).

95. Halley, M. C., Halverson, C. M. E., Tabor, H. K. & Goldenberg, A. J. Rare Disease, Advocacy and Justice: Intersecting Disparities in Research and Clinical Care. Am. J. Bioeth. 23, 17–26 (2023).

96. Kingdom, R. & Wright, C. F. Incomplete Penetrance and Variable Expressivity: From Clinical Studies to Population Cohorts. Front. Genet. 13, (2022).

97. Cutler, D. J., Jodeiry, K., Bass, A. J. & Epstein, M. P. The Quantitative Genetics of Human Disease: 2 Polygenic Risk Scores. Human Population Genetics and Genomics 1–65 (2024) doi:10.47248/hpgg2404030008.

98. Weiner, D. J. et al. Polygenic architecture of rare coding variation across 394,783 exomes. Nature 614, 492–499 (2023).

99. Carrasquillo, M. M. et al. Genome-wide association study and mouse model identify interaction between RET and EDNRB pathways in Hirschsprung disease. Nat. Genet. 32, 237–244 (2002).

100. Simons, Y. B., Turchin, M. C., Pritchard, J. K. & Sella, G. The deleterious mutation load is insensitive to recent population history. Nat. Genet. 46, 220–224 (2014).

101. Frazer, K. A., Murray, S. S., Schork, N. J. & Topol, E. J. Human genetic variation and its contribution to complex traits. Nat. Rev. Genet. 10, 241–251 (2009).

102. Kasowski, M. et al. Variation in Transcription Factor Binding Among Humans. Science (1979). 328, 232–235 (2010).

