## Supplementary Files and Tables for "Population Differences in the Epidemiology, Phenotype, and Genetics of Hirschsprung Disease in the United States"

**Figure S1: Comparison of self-identified race and ethnicity (SIRE) with genetic ancestry and ancestry-inferred population in 205 HDRC individuals.**

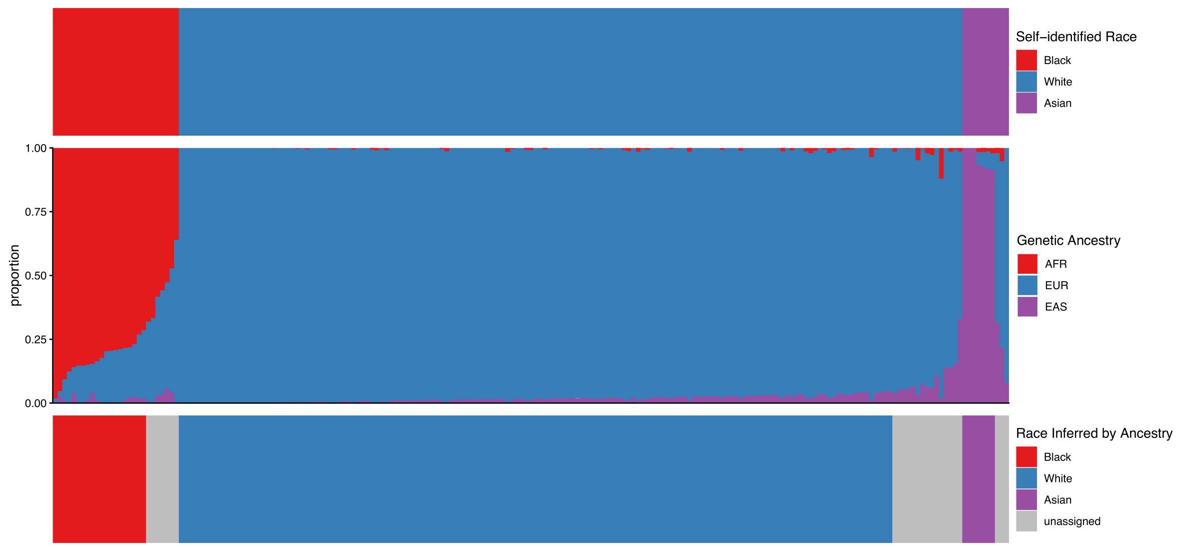


Top: self-identified race and ethnicity (SIRE) from the HDRC questionnaire.
Middle: estimated genetic ancestry of the corresponding individuals.
Bottom: ancestry-inferred race and ethnicity based on relative components of genetic ancestry.

**Figure S2: Geographic distribution of non-Hispanic Black and non-Hispanic White HSCR patients recruited in the HDRC.**

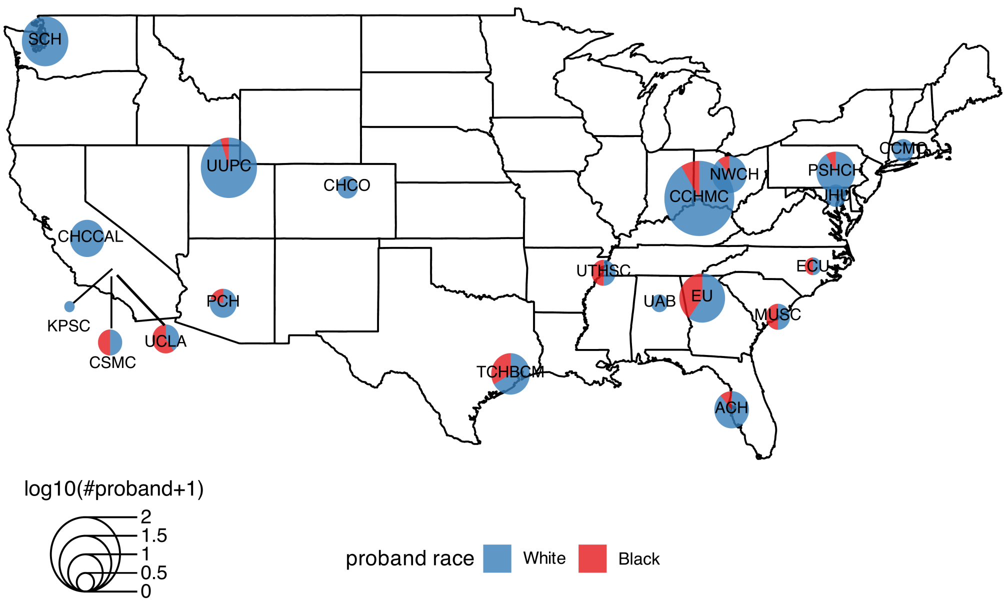

**Figure S3: Population-specific incidence of HSCR in the NBDPN, Epic-Cosmos and the literature.**

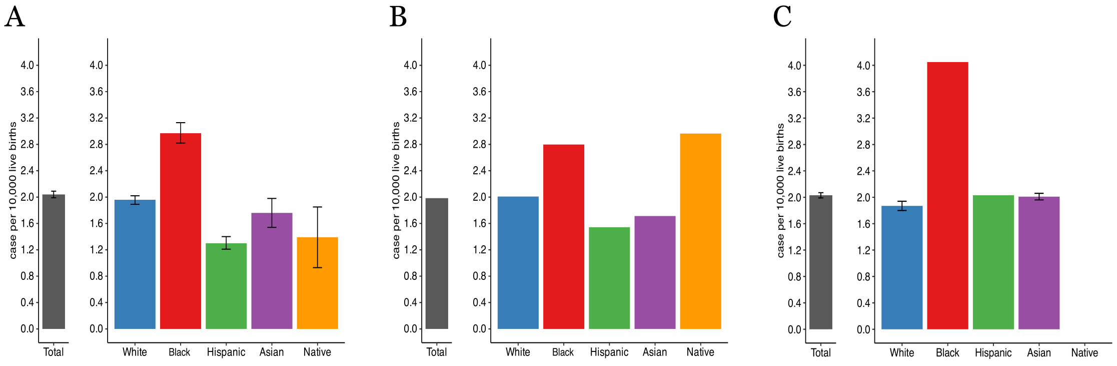


(A) U.S. HSCR incidence from the National Birth Defects Prevention Networks (NBDPN) data for the years 1996-2010. Information was extracted from 35 state-level surveillance data on 30,236,567 individuals.
(B) U.S. HSCR incidence from Epic Cosmos for the years 1996-2025. Data were obtained from electronic health records of >190 hospitals covering >7 million individuals. Incident cases were defined as individuals diagnosed with HSCR by 3 years of age.
(C) HSCR incidence across the world from a literature review with mid-study periods of 1996. Data were extracted from 9 studies covering 49,667,555 individuals.

**Figure S4: Clinical features of HSCR patients by SIRE in the HDRC study.**

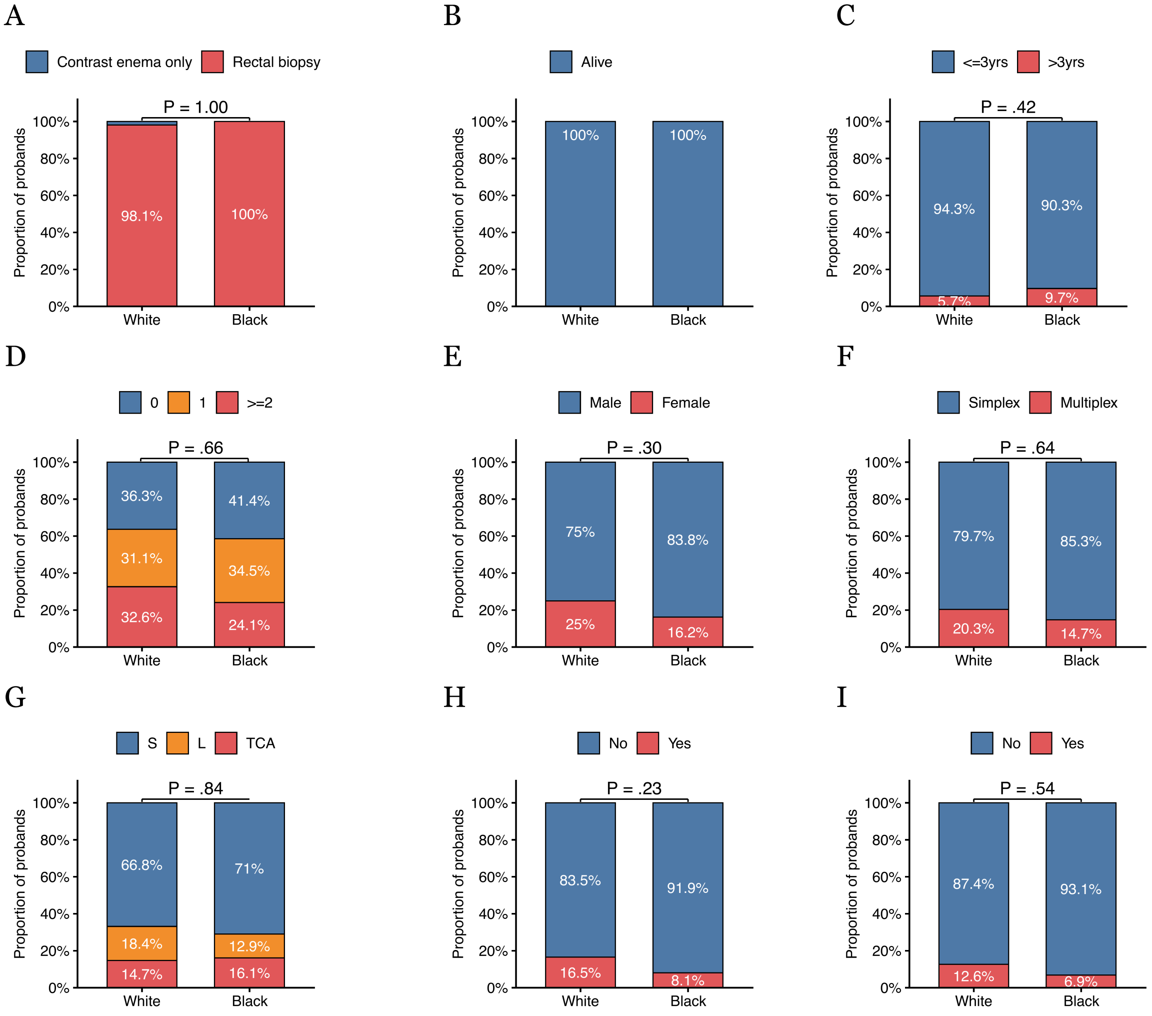


Phenotypic classifications of 237 White and 37 Black HSCR probands from the HDRC study by (A) diagnostic test, (B) mortality as of 2025, (C) age at diagnosis, (D) disease severity (higher score: higher severity), (E) sex, (F) familiality, (G) segment length of aganglionosis, (H) presence of additional HSCR related congenital anomalies, and (I) Down syndrome status.

**Table S1: Numbers of states and time periods of HSCR surveillance in the NBDPN.**

| Period | # States reporting on HSCR | Self-described race/ethnicity |
|---|---|---|
| 1989 – 1995* | 13 | White,<br>Other |
| 1996 - 2000 | 20 | White (non-Hispanic),<br>Black (non-Hispanic),<br>Hispanic,<br>Asian or Pacific Islander (non-Hispanic),<br>American Indian or Alaska Native (non-Hispanic) |
| 2001 - 2005 | 22 | White (non-Hispanic),<br>Black (non-Hispanic),<br>Hispanic,<br>Asian or Pacific Islander (non-Hispanic),<br>American Indian or Alaska Native (non-Hispanic) |
| 2006 - 2010 | 32 | White (non-Hispanic),<br>Black (non-Hispanic),<br>Hispanic,<br>Asian or Pacific Islander (non-Hispanic),<br>American Indian or Alaska Native (non-Hispanic) |

*The self-described race/ethnicity categories were different in the period of 1989 – 1995 as compared to others.

**Table S2: Studies used for literature review analysis of HSCR incidence**

| Study name | Mid-study period | Study area | Region |
|---|---|---|---|
| Taguchi, 2017 | 1996 | Japan | East Asia |
| Löf Granström, 2016 | 1998 | Sweden | Europe |
| Singh, 2003 | 1999 | Australia | Oceania |
| Best, 2012 | 2000 | North of England | Europe |
| Nasr, 2017 | 2003 | Ontario, Canada | North America |
| Chia, 2016 | 2005 | Taiwan | East Asia |
| Anderson, 2018 | 2005 | California, USA | North America |
| Taghavi, 2019 | 2008 | New Zealand | Oceania |
| Bradnock, 2017 | 2012 | UK and Ireland | Europe |

**Table S3: Major medical institutions and clinics participating in the Hirschsprung Disease Research Consortium (HDRC) during 2011-2024.**

| Study name | Institution | Country | State/Province | Zip/Post code |
|---|---|---|---|---|
| HDRCWCH | Winnipeg Children's Hospital | Canada | Manitoba | R3E 0Z3 |
| HDRCHSC | Hospital for Sick Children | Canada | Ontario | M5G 1X8 |
| HDRCNAD | Gadja Mada University, Indonesia | Indonesia | Special Region of Yogyakarta | 55281 |
| HDRCUAB | University of Alabama at Birmingham | US | AL | 35233 |
| HDRCPCH | Phoenix Children's Hospital | US | AZ | 85016 |
| HDRCCHCCAL | Valley Children's Hospital | US | CA | 93636 |
| HDRCCSMC | Cedars-Sinai Med Center | US | CA | 90048 |
| HDRCKPSC | Kaiser Permanente SC | US | CA | 90027 |
| HDRCUCD | University of California, Davis | US | CA | 95817 |
| HDRCUCLA | University of California, Los Angeles | US | CA | 90095 |
| HDRCCHCO | Children's Hospital Colorado | US | CO | 80045 |
| HDRCCCMC | Connecticut Children's | US | CT | 6106 |
| HDRCACH | All Children's Hospital | US | FL | 33701 |
| HDRCEU | Emory/Children's Atlanta | US | GA | 30322 |
| HDRCJHU | Johns Hopkins University | US | MD | 21287 |
| HDRCUMICH | University of Michigan | US | MI | 48109 |
| HDRCECU | East Carolina University | US | NC | 27858 |
| HDRCCCHMC | Cincinnati Children's Hospital Medical Center | US | OH | 45229 |
| HDRCNWCH | Nationwide Children's Hospital | US | OH | 43205 |
| HDRCPSHCH | Penn State Hershey Children's Hospital | US | PA | 17033 |
| HDRCMUSC | Medical Univ of SC | US | SC | 29425 |
| HDRCUTHSC | University of Tennessee Health Science Center | US | TN | 38105 |
| HDRCTCHBCM | Texas Children's/ Baylor College of Medicine | US | TX | 77030 |
| HDRCUUPC | University of Utah | US | UT | 84108 |
| HDRCSCH | Seattle Children's Hospital | US | WA | 98145 |

**Table S4: HSCR incidence ratios across populations in different surveillance time intervals from the NBDPN data.**

| | **1996-2000** | **2001-2005** | **2006-2010** |
|---|---|---|---|
| **Black vs. White** | 1.60 (1.42-1.8) $p< 0.0001$ | 1.46 (1.31-1.62) $p< 0.0001$ | 1.48 (1.34-1.64) $p< 0.0001$ |
| **Hispanic vs. White** | 0.68 (0.59-0.80) $p< 0.0001$ | 0.61 (0.53-0.69) $p< 0.0001$ | 0.70 (0.61-0.79) $p< 0.0001$ |
| **Asian vs. White** | 1.04 (0.81-1.33) $p=0.77$ | 0.90 (0.72-1.11) $p=0.30$ | 0.79 (0.64-0.98) $p=0.034$ |
| **Native American vs. White** | 0.73 (0.38-1.40) $p=0.33$ | 1.12 (0.71-1.76) $p=0.63$ | 1.22 (0.86-1.75) $p=0.26$ |

Each cell presents the incidence rate ratio, 95% confidence interval and P value per time interval.

**Table S5: Multivariate regression analysis of HSCR incidence with respect to SIRE, study period and geographic locations from NBDPN data.**

| Variables | β | P |
|---|---|---|
| **Black (vs White)** | 0.90 | $2.1x10^{-5}$ |
| **Hispanic (vs White)** | -0.43 | 0.04 |
| **Asian (vs White)** | -0.21 | 0.34 |
| **Native American (vs White)** | -0.02 | 0.94 |
| **Time Interval** | -0.02 | 0.20 |
| **Longitude** | 0.01 | 0.44 |
| **Latitude** | -0.02 | 0.47 |

Multivariate regression model was performed using the R GLM package by setting HSCR incidence as response variable and using SIRE, study time period, longitude and latitude as independent variables. Ancestry was studied using Whites as the reference group; all other explanatory variables were numeric.

**Table S6: Surveillance sample sizes for the NBDPN, Epic Cosmos and literature review data.**

| Population | NBDPN | Epic Cosmos | Literature Review |
|---|---|---|---|
| Total | 30,236,567 | 7,573,207 | 49,667,555 |
| White | 17,408,132 | 3,903,473 | 13,167,548 |
| Black | 4,841,311 | 1,283,465 | 656,790 |
| Hispanic | 5,954,922 | 1,257,922 | 4,896,552 |
| Asian | 1,397,875 | 356,105 | 29,099,326 |
| Native American | 278,062 | 101,183 | - |
| Other/Unknown | 356,265 | 671,059 | 2,000,000 |

**Table S7: Comparisons of rare pathogenic coding variant burden across the core 24 HSCR risk genes in White and Black patients from the HDRC study.**

| | PA Type | White (N, %) | African American (N, %) | P |
|---|---|---|---|---|
| *RET* | LoF | 11 (5.4%) | 2 (6.1%) | 0.69 |
| | missence | 15 (7.4%) | 2 (6.1%) | 1 |
| | All | 27 (13.2%) | 4 (12.1%) | 1 |
| *EDNRB* | LoF | 3 (1.5%) | 0 (0%) | 1 |
| | missence | 3 (1.5%) | 1 (0%) | 0.45 |
| | All | 6 (2.9%) | 1 (3.0%) | 1 |
| All 24 risk genes combined | LoF | 19 (9.3%) | 2 (6.1%) | 0.75 |
| | missence | 38 (18.6%) | 4 (12.1%) | 0.47 |
| | All | 54 (26.5%) | 6 (18.2%) | 0.39 |

Coding variants (All) were further grouped into missense and loss-of-function (LoF) by variant class from 204 White and 33 Black HSCR patients. The table indicates the number and percent of probands in each cell considering *RET*, *EDNRB* and all 24 HSCR risk genes combined. P values were calculated using Fisher's exact tests.

**Table S8: Comparisons of 5 risk enhancer variant haplotypes at the *RET* locus by population.**

| Haplotype | # Risk alleles | Black/African American | | | | White | | | |
|---|---|---|---|---|---|---|---|---|---|
| | | *case freq* | *ctrl freq* | *OR (95% CI)* | *P* | *case freq* | *ctrl freq* | *OR (95% CI)* | *P* |
| CTGAC | 0 | 0.62 | 0.81 | (1=ref) | — | 0.22 | 0.51 | (1=ref) | — |
| CTGA**T** | 1 | 0.08 | 0.02 | **6.34 (1.63–24.63)** | **$1.06\times10^{-2}$** | 0.2 | 0.09 | **5.05 (3.59–7.1)** | **$1.64\times10^{-19}$** |
| **GCA**AC | 3 | 0.08 | 0.02 | 4.23 | 0.028 | — | — | — | — |
| **GCAG**C | 4 | 0.05 | 0.12 | 0.51 | 0.42 | 0.12 | 0.25 | 1.11 | 0.57 |
| **GCAGT** | 5 | 0.15 | 0.04 | **5.07 (1.99–12.97)** | **$1.07\times10^{-3}$** | 0.44 | 0.15 | **6.94 (5.22–9.23)** | **$2.84\times10^{-43}$** |

Observed frequencies of haplotypes and counts (N) in cases and controls, together with the odds ratio (with respect to the reference haplotype containing the fewest risk alleles), are shown for 33 Black/African American cases, 204 White cases, 129 Black/African American controls and 970 White controls. Risk alleles are bolded. P values were calculated using Fisher's exact test, and significant ones are bolded. The 5 risk enhancer variants for each haplotype are in order: rs788263 |rs788261 |rs788260 |rs2506030 |rs2435357.

**Table S9: Counts of common non-coding variants within the expression domain for each of the 24 core HSCR risk genes in Whites and Blacks.**

| | Enrichment | | | |
|---|---|---|---|---|
| **Gene** | **none** | **both** | **Whites** | **Blacks** |
| *ACSS2* | 3 | | 1 | |
| *ADAMTS17* | 1 | | | |
| *EDN3* | 5 | | 3 | |
| *EDNRB* | 13 | | | |
| *ELP1* | 3 | | 2 | |
| *ENO3* | 4 | | 3 | |
| *GDNF* | 2 | | | |
| *GFRA1* | 3 | | | |
| *KIFBP* | | | 1 | |
| *NRG1* | 3 | | | |
| *NRTN* | 2 | | 1 | |
| *PRXL2A* | | | 1 | |
| *RET* | 280 | 28 | 1 | 1 |
| *SEMA3C* | 2 | | | |
| *SEMA3D* | 2 | | | |
| *SH3PXD2A* | 1 | | | |
| *SLC27A4* | 1 | | 1 | |
| *SOX10* | | | | 3 |
| *ZEB2* | 1 | | | |

*Enrichment was defined as 2-fold or greater increase in the coded allele frequency in cases versus controls with at least marginal significance.
Case allele frequency was calculated separately from 204 White and 33 Black patients in the HDRC study. Control allele frequency was obtained from gnomAD African American or European (non-Finnish) samples. P values were calculated using Fisher's Exact test with significance corrected as 0.05/ number of variants per gene. Marginal significance was defined as 2 times significance.
