## Supplementary Methods for "Population Differences in the Epidemiology, Phenotype, and Genetics of Hirschsprung Disease in the United States"

### Global Literature Review of HSCR Incidence

To contextualize U.S. HSCR incidence estimates within the global literature, a systematic search of PubMed was conducted using the terms "(Hirschsprung OR megacolon) AND (incidence OR prevalence)", restricted to English-language publications from 1950 onwards — the period following establishment of pathology-based HSCR diagnosis. A total of 361 records were identified; no duplicates were detected after deduplication. After title and abstract screening, 282 records were excluded for not reporting HSCR incidence or birth prevalence, leaving 79 articles for full-text review. These articles, together with references identified through citation tracking, were reviewed in full.

At the full-text stage, studies were retained if they satisfied all of the following criteria: (1) explicitly reported HSCR incidence or birth prevalence as a quantitative numeric outcome; (2) were based on population-level surveillance systems or multicenter medical records spanning a defined geographic region, rather than single referral centers; (3) clearly specified the geographic scope of the denominator population (country, state/province, or region) screened; (4) provided explicit start and end years of case ascertainment; and (5) reported or allowed derivation of both the number of HSCR cases and the reference population number (live births or total population at risk). Studies were excluded at the full-text stage if they lacked explicit case counts or live-birth denominators, failed to define the study population or period, or were based on a single care center or referral-based sample with limited population generalizability.

Of the 79 full-text articles, 31 explicitly reported quantitative HSCR incidence or birth prevalence estimates (Supplementary File1). Among them, 15 were further excluded for relying on single-center data, lacking explicit case counts or live-birth denominators, or failing to define the study population or period. The remaining 16 population-based or multicenter studies spanning North America, Oceania, Europe, and East Asia were included. Of these, 9 studies with mid-study periods approximating 1996 were sub-selected in downstream analysis for temporal consistency with the National Birth Defects Prevention Network (NBDPN). The literature review flow is summarized below.

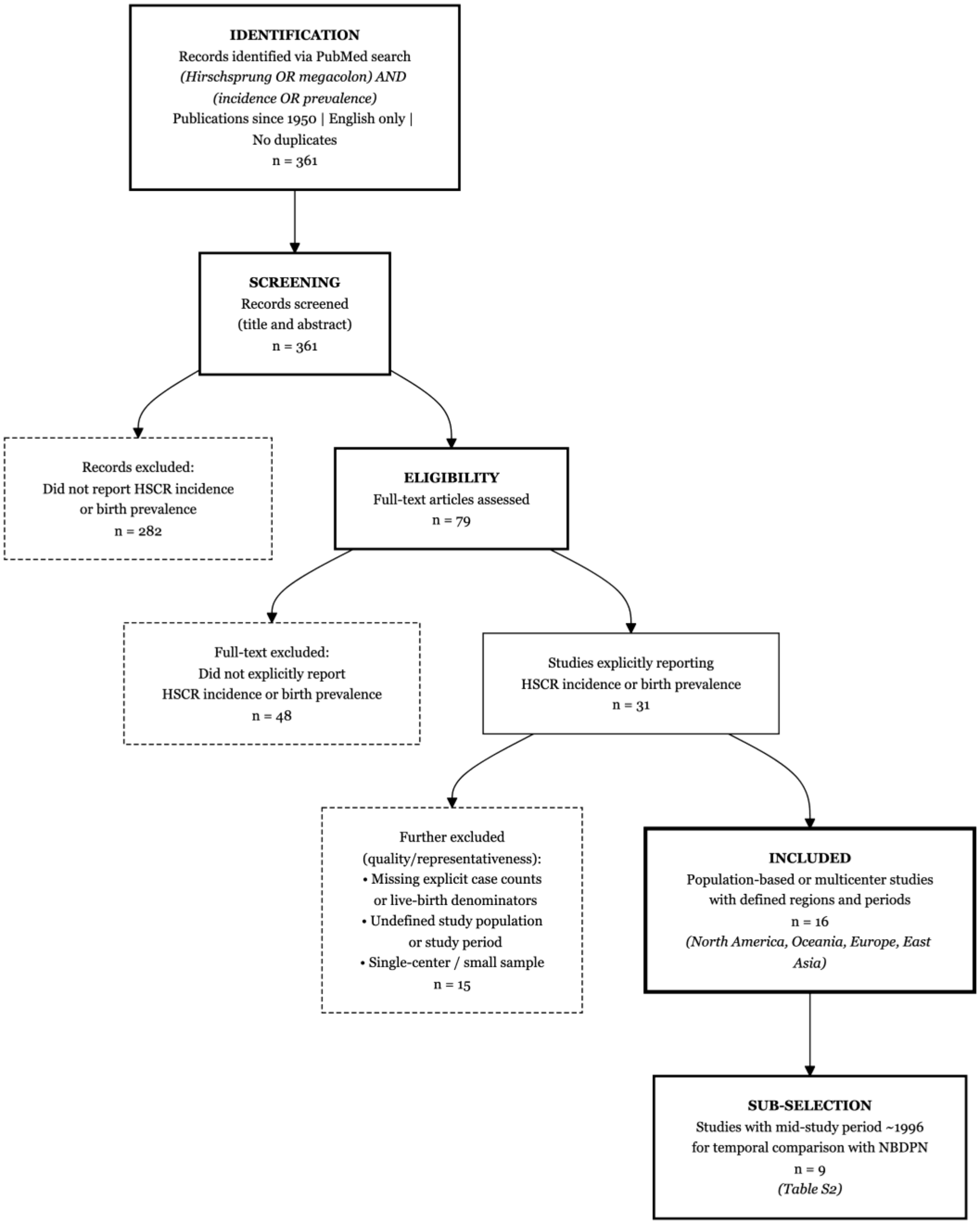
IDENTIFICATION
Records identified via PubMed search
(Hirschsprung OR megacolon) AND
(incidence OR prevalence)
Publications since 1950 | English only |
No duplicates
n = 361
SCREENING
Records screened
(title and abstract)
n = 361
Records excluded:
Did not report HSCR incidence
or birth prevalence
n = 282
ELIGIBILITY
Full-text articles assessed
n = 79
Full-text excluded:
Did not explicitly report
HSCR incidence or birth prevalence
n = 48
Studies explicitly reporting
HSCR incidence or birth prevalence
n = 31
Further excluded
(quality/representativeness):
• Missing explicit case counts
or live-birth denominators
• Undefined study population
or study period
• Single-center / small sample
n = 15
INCLUDED
Population-based or multicenter studies
with defined regions and periods
n = 16
(North America, Oceania, Europe, East
Asia)
SUB-SELECTION
Studies with mid-study period ~1996
for temporal comparison with NBDPN
n = 9
(Table S2)

### DNA Extraction and Quality Assessment

Genomic DNA was extracted from 2,366 blood samples and 117 saliva samples of HSCR probands and their relatives from the HDRC cohort. DNA from blood and saliva samples was extracted using the Gentra Puregene Kit (Qiagen; catalog no. 69504) and the Oragene-DNA Kit (DNA Genotek; catalog no. OG-600), respectively, following the manufacturers' protocols. DNA concentration was measured using the PicoGreen assay (Thermo Fisher Scientific; catalog # P11496) and DNA quality was assessed using NanoDrop spectrophotometry (Thermo Fisher Scientific; model T042) by evaluating the 260/280 absorbance ratio. For blood-derived DNA, concentrations ranged from 157.7 to 795.4 ng/µL, with 260/280 ratios ranging from 1.79 to 1.89. For saliva-derived DNA, concentrations ranged from 64.0 to 503.4 ng/µL, with 260/280 ratios ranging from 1.63 to 1.96, indicating acceptable DNA quality for genomic sequencing. Following quality control, DNA samples were aliquoted and plated on 96-well plates. Each sample was diluted with TE buffer (Invitrogen; catalog no. AM9858) to a final concentration of 15 ng/µL in a total volume of 50 µL. which were then shipped to the Broad Institute for whole-genome sequencing (WGS).

### Whole-Genome Sequencing (WGS) and Variant Calling Pipeline

WGS of our samples was funded by the NIH Gabriella Miller Kids First Pediatric Research Program (project number HD110884-01) and performed at the Broad Institute using a centralized, automated sequencing and analysis pipeline from our prepared DNA samples. Over 90% of the DNA samples (n=2,132) that passed the Broad's multiple quality control steps, including assessments of DNA integrity and quantity, advanced to WGS. Sequencing libraries were prepared using a PCR-free WGS protocol (KAPA HyperPrep Kit catalog no. 07962363001) and sequenced on Illumina platforms using paired-end 150 base pair reads, targeting a mean genome-wide coverage of 30x. Sequencing reads for each sample were aligned to the human reference genome (hg38) and consolidated into a single CRAM file per sample.

Variant discovery was performed by the Broad using the Genome Analysis Toolkit version 4 (GATK4)[1] best-practices pipeline for germline short-variant discovery. Briefly, aligned CRAM files were processed using HaplotypeCaller to generate individual, interval-scattered genomic VCF (gVCF) files. Joint variant calling across all samples was then performed using the Genomic Variant Store (GVS) pipeline, which enables scalable, interval-based joint genotyping. All detected variants were retained in the final call set along with their corresponding quality and filter annotations in interval-scattered, joint-called vcf.gz format.

The Broad Institute returned individual CRAM files and interval-scattered, joint-called VCF.gz files, which were deposited in the NIH's data portal for secure data transfer. Joint-called VCF files were downloaded and used for all downstream analyses. For this study, unrelated HDRC probands were extracted from the master joint-called VCF files using bcftools.[2]

### Quality Control and Variant Filtering

Variant-level and sample-level quality control (QC) was performed using a standardized pipeline following GATK's best practices and previously published WGS protocols.[3] Briefly, only variants that passed GATK's Variant Quality Score Recalibration (VQSR) filters were retained. Analyses were restricted to bi-allelic variants. Additional hard filters were applied, including a minimum read depth (DP) of 10, genotype quality (GQ) of at least 20, and allelic balance (AB) between 0.20 and 0.80 for heterozygous genotypes. Variants located within low-complexity regions (LCRs) were excluded. Variants and samples with greater than 10% missingness were removed.

Additional sample-level QC was performed to detect and remove outliers using a graph-based approach based on 13 quality metrics derived from GATK's CollectVariantCallingMetrics. These metrics included the heterozygous-to-homozygous variant ratio, percentage of genotypes with GQ of zero, total number of single-nucleotide variants (SNP), percentage of variants present in dbSNP, percentage of multiallelic variants, SNP reference bias, novel insertion-to-deletion ratio, novel transition-to-transversion ratio, dbSNP insertion-to-deletion ratio, dbSNP transition-to-transversion ratio, total number of insertions and deletions, number of singleton variants, and percentage of dbSNP insertions and deletions. 0.6% samples identified as outliers were excluded from downstream analyses

**Ancestry Estimation**

Genetic ancestry of the unrelated HDRC probands were estimated by mapping and comparing variants in the proband samples with variants in the 1000G reference samples,[4] using only common (minor allele frequency, MAF > 10%), linkage disequilibrium (LD)-pruned ($r2 < 0.3$), bi-allelic, autosomal variants present in both HDRC probands and 1000G samples, using the software ADMIXTURE.[5] Probands were clustered based on their relative proportions of European (EUR), African (AFR) and East Asian (EAS) genetic ancestries.

**Variant Annotation and Pathogenic Coding Variant Definition**

Variants were functionally annotated using the Ensembl Variant Effect Predictor (VEP).[6] Additional pathogenicity annotations were applied to protein coding variants using tools of phylop241way,[7] VEST4,[8] LOFTEE,[9] spliceAI,[10] REVEL,[11] and metaRNN,[12] based on variants' predicated protein impact and conservation.

Pathogenic coding variants were defined and prioritized using the following criteria: missense variants with a REVEL score greater than 0.5; stop-gain and frameshift variants classified as high confidence (HC) by LOFTEE; splice donor or acceptor variants classified as HC by LOFTEE or with a SpliceAI score greater than 0.8; and insertions or deletions (INDELs) with a VEST4 score greater than 0.5, metaRNN score greater than 0.5, or phyloP241way score greater than 6. These variants were further restricted to rare variants, defined as those with a global allele frequency less than 1% in the gnomAD version 4 reference population.[13] Only variants meeting both rarity and pathogenicity criteria were included in downstream rare coding pathogenic variant burden analyses.

### Supplementary References


1. GATK Team. Genotype Refinement workflow for germline short variants. https://gatk.broadinstitute.org/hc/en-us/articles/360035531432-Genotype-Refinement-workflow-for-germline-short-variants (2024).

2. Danecek, P. *et al.* Twelve years of SAMtools and BCFtools. *Gigascience* **10**, (2021).

3. Sealock, J. M. *et al.* Tutorial: guidelines for quality filtering of whole-exome and whole-genome sequencing data for population-scale association analyses. *Nat. Protoc.* **20**, 2372–2382 (2025).

4. Byrska-Bishop, M., Evani, U. S., Zhao, X. & Basile, A. O. High coverage whole genome sequencing of the expanded 1000 Genomes Project cohort including 602 trios. *bioRxiv* 2021.02.06.430068 (2021).

5. Alexander, D. H. & Lange, K. Enhancements to the ADMIXTURE algorithm for individual ancestry estimation. *BMC Bioinformatics* **12**, 246 (2011).

6. McLaren, W. *et al.* The Ensembl Variant Effect Predictor. *Genome Biol.* **17**, 122 (2016).

7. Zoonomia. A comparative genomics multitool for scientific discovery and conservation. *Nature* **587**, 240–245 (2020).

8. Douville, C. *et al.* Assessing the Pathogenicity of Insertion and Deletion Variants with the Variant Effect Scoring Tool (VEST-Indel). *Hum. Mutat.* **37**, 28–35 (2016).

9. https://github.com/konradjk/loftee. LOFTEE.

10. Jaganathan, K. *et al.* Predicting Splicing from Primary Sequence with Deep Learning. *Cell* **176**, 535-548.e24 (2019).

11. Ioannidis, N. M. *et al.* REVEL: An Ensemble Method for Predicting the Pathogenicity of Rare Missense Variants. *The American Journal of Human Genetics* **99**, 877–885 (2016).

12. Li, C., Zhi, D., Wang, K. & Liu, X. MetaRNN: differentiating rare pathogenic and rare benign missense SNVs and InDels using deep learning. *Genome Med.* **14**, 1–14 (2022).

13. Gudmundsson, S. *et al.* Variant interpretation using population databases: Lessons from gnomAD. *Hum. Mutat.* **43**, 1012–1030 (2022).
